# Functional Variant Discovery identifies a novel genetic link between SPRY2, wood smoke, and asthma

**DOI:** 10.1101/2025.06.20.25330004

**Authors:** Arnav Gupta, Amber Dahlin, Alejandra Macario, Fabienne Gally, Michael R. Weaver, Samuel Guarino, Louisa Kahn, Lynn Sanford, Margaret A. Gruca, Michael H. Cho, Robin D. Dowell, Scott T. Weiss, Sarah K. Sasse, Anthony N. Gerber

## Abstract

As a consequence of climate change and land use policies, there has been a historic rise in wildfire smoke across the United States and the world. While the deleterious effects of wildfire smoke and associated air pollution on asthma outcomes are established epidemiologically, genetic risks and molecular mechanisms of how wildfire smoke affects asthma are unknown. This knowledge gap hinders the identification of high-risk individuals and the creation of targeted therapies or recommendations to protect these individuals. We identified 52 genetic risk variants that colocalized with genomic responses to wood smoke particles (WSP), a model of wildfire particulate matter, and associated with asthma in the Genetic Epidemiology Research on Aging (GERA) cohort. We used additional filters to prioritize variants for direct testing of allele-dependent transcriptional regulatory function in plasmid reporters. We found that the rs3861144 variant (Odds Ratio_asthma_ = 1.036) changes *SPRY2* responses to WSP in airway epithelial cells, which are involved in Interleukin-8 secretion, Extracellular Signal-related Kinase (ERK) activation, and mechanical scratch repair in cell culture. These findings provide insights into the molecular pathways through which WSP may influence asthma risk and propose genetic candidates that warrant further study for their potential as clinical tools for asthma.

## Introduction

Climate change is contributing to a historic rise in wildfire smoke and reversing decades of improvements in air quality made by environmental policy (1–3). The scope of this problem is extending across North America and the world, increasing the population exposed to wildfire air pollution (4). Wildfire smoke is difficult to regulate or prevent. Climate policy and wildland management strategies will take decades to reverse the trend of expanding wildfires, and deteriorating air quality is expected over the next 30 years (5). It is therefore critical to understand the health consequences of wildfire air pollution and implement detailed strategies to manage and mitigate this developing healthcare crisis.

The profound health impacts of wildfire smoke and air pollution are well established at the population scale (6,7) and the impact on respiratory diseases, such as asthma, is a major concern. Wildfire smoke increases levels of ambient air pollutants, such as particulate matter less than 2.5 um (PM2.5), and causes morbid asthma outcomes, such as loss of asthma control, emergency room visits and hospitalizations (8–10). Data suggest ambient PM2.5 may also contribute to reduced lung function and the development of asthma (11–13). These major concerns highlight critical knowledge gaps in our current understanding of how PM2.5 from wildfire smoke impacts respiratory biology and asthma. First, the molecular genetic mechanisms that connect wildfire PM2.5 with asthma have not been identified, preventing the development of targeted treatments that mitigate asthma development or exacerbation. Second, there is significant variation in how individuals respond to airborne exposures, such as PM2.5 or wildfire smoke (14), and public health guidelines, which only advise exposure avoidance, have limited efficacy, as restricting large population segments from outdoor activity for weeks during wildfire events is impractical. A better understanding of how PM2.5 and wildfire air pollution contributes to asthma risk and identification of high-risk individuals are critical to address this growing problem.

Genetic analysis is a powerful tool to inform disease mechanisms, define individual risk, and advance our understanding of the relationships between PM2.5, wildfire smoke, and asthma. However, there is a significant gap between the effect sizes of established genetic variants and asthma heritability, which is estimated at up to 70% of causation in certain studies (15). This suggests that additional asthma risk factors that are not captured in genome-wide association studies (GWAS), such as environmental exposures, can influence genetic effects. This concept is supported by discoveries of gene mechanisms that connect air pollution exposure with asthma, such as mutations regulating *NQO1*, *GSTM1* and *GSTP1* (16–18). Identification of additional conditional genetic risk variants, which primarily contribute to asthma risk in the presence of exposure, has the potential to define novel asthma mechanisms associated with wildfire PM2.5 and individuals with high risk.

Several challenges limit the identification of exposure-conditional risk variants, despite the major advances achieved by asthma genetic studies. First, integrating environmental variables in GWAS is highly challenging due to a myriad of reasons, including exposure variability, duration of exposure effects, and types of exposures, among others. Genetic-environment interaction studies have approached this challenge by quantifying environmental variables through geolocalization and retrospective measurements of local air quality (19), but these studies are limited by the precision of geolocalization, resolution of air pollutant measurements, and difficulty with compounding exposures based on where study subjects have lived over decades. Second, linking specific genetic variants with cell and molecular functions can be challenging. GWAS lead variants often reside in non-coding regions (20) and represent statistical but not functional association priorities. Tools, such as functional scoring and expression quantitative trait locus (eQTL) analysis, can improve associations between GWAS variants and gene regulation but fail to leverage the full power of GWAS results, which may contain additional functional variants that do not meet statistical thresholds. These tools also do not account for the effects of relevant environmental exposures. Therefore, novel approaches that build on top of existing GWAS data, incorporate environmental exposures, and directly connect variants with function are necessary to realize the full potential of genetic analysis.

In this study, we applied our novel approach to variant discovery (21) to identify functional genetic variants and molecular mechanisms that connect a wildfire PM2.5 model with asthma risk. Using wood smoke particles (WSP) (22,23) to model wildfire PM2.5, we identified and biologically validated a set of exposure-conditional genetic variants using population data of lung function, transcription factor binding motifs, and epigenetic features of live airway epithelia obtained from bronchoscopy. We directly tested the potential for high-priority variants to change transcriptional function, connected variants with gene targets, and tested the function of these target genes in airway epithelial cell biology. Our results were highlighted by the discovery of rs3861144, a functional variant that changes *SPRY2* responses to WSP exposure and potentially contributes to asthma through cytokine secretion and injury-repair programs.

## Methods and Materials

Additional methods are provided in the Online Data Supplement due to word limit constraints.

### Genetic discovery data set and Analysis

Genome-wide and phenotype data were utilized from the Genetic Epidemiologic Research on Aging (GERA) study, a large, multi-ethnic asthma cohort including over 110,000 subjects (68,623 total asthma cases across four ethnic groups) with extensive electronic medical records and genome-wide SNP genotypes (>8 million typed and imputed markers). We previously reported multiple novel associations with asthma in four major ethnic groups in GERA (24). The current analysis was limited to the ethnic subset of the cohort with the highest percentage of asthma cases and therefore the greatest statistical power; the final data set included information from 16,274 non-Hispanic white asthmatic cases and 38,269 controls (54,543 total samples). We applied methods previously developed by our group (21) to regulatory elements identified in WSP-exposed Beas-2B cells by Precision Run-on sequencing (PRO-seq) (n=18,552) (25). SNPs that overlapped regions within 50 bp (+/-) of regulatory element centers were selected for association analysis. Tests for association of additive genotype with asthma status (dichotomized outcome variable) were conducted using multivariate logistic regression models, adjusted for covariates, and the significance of associations was determined using a max(T) permutation-based approach with 5000 permutations and a false-discovery rate (FDR) threshold of 0.05. All analyses were performed using R statistical computing software and PLINK 1.9.

### CRISPR-Cas9 RNP transfection and nucleofection

We performed genomic deletion of the transcriptional regulatory element (TRE) containing rs3861144 and *SPRY2* using the Alt-R CRISPR-Cas9 system supplied and guided by IDT. Custom-designed crRNAs, which target two sites within the TRE or the *SPRY2* gene body, and a negative control crRNA (IDT) were duplexed (5 min at 95C) with tracrRNAs and then complexed with Cas9 enzyme (1:1 molar ratio of duplex to Cas9) to a final concentration of 1 uM. This ribonucleic protein complex was transfected into Beas-2B cells using Lipofectamine CRISPRmax (Life Technologies) or electroporated into primary small airway epithelial cells (SAECs) using the Lonza nucleofector IIb system and the Basic Kit for Epithelial Cells. Nucleofection protocol was adapted from a previous study (26) for the I/IIb system using the T-020 program. Genome editing was confirmed by PCR amplification of genomic DNA (DNeasy Blood and Tissue kit – Qiagen) or Western Blot. Guide RNA sequences are provided in Supplemental Table E2.

## Results

### Identification of functional variant candidates associated with asthma risk and wood smoke particle exposure

To connect airway epithelial wood smoke particle (WSP) responses with mechanisms of asthma, we re-analyzed nascent transcription profiles from an airway epithelial cell line (Beas-2B) exposed to WSP for 30 or 120 minutes (25). Within 18,552 dynamic transcriptional regulatory element (TRE, transcriptional start sites excluded) responses to WSP (100 bp regions, increased or decreased transcription between vehicle/control and WSP at 30 or 120 minutes), we tested 4373 single nucleotide polymorphisms (SNPs, minor allele frequency > 0.05) and identified 52 candidates associated with asthma phenotype in the non-Hispanic-white subpopulation of the GERA cohort (False Discovery Rate, FDR < 0.05, max(T) permutation), which are designated “WSP-SNPs” in the remainder of this manuscript (Figure 1A). These candidates predominantly reside in non-coding genomic regions (Figure 1B). A representative example of a WSP-SNP, rs28362965, is depicted in Figure 1C. Here, WSP exposure increases transcription of both the TRE containing rs28362965 and *HK2.* A full list of the 52 WSP-SNPs is provided in Table 1.

**Figure 1.**
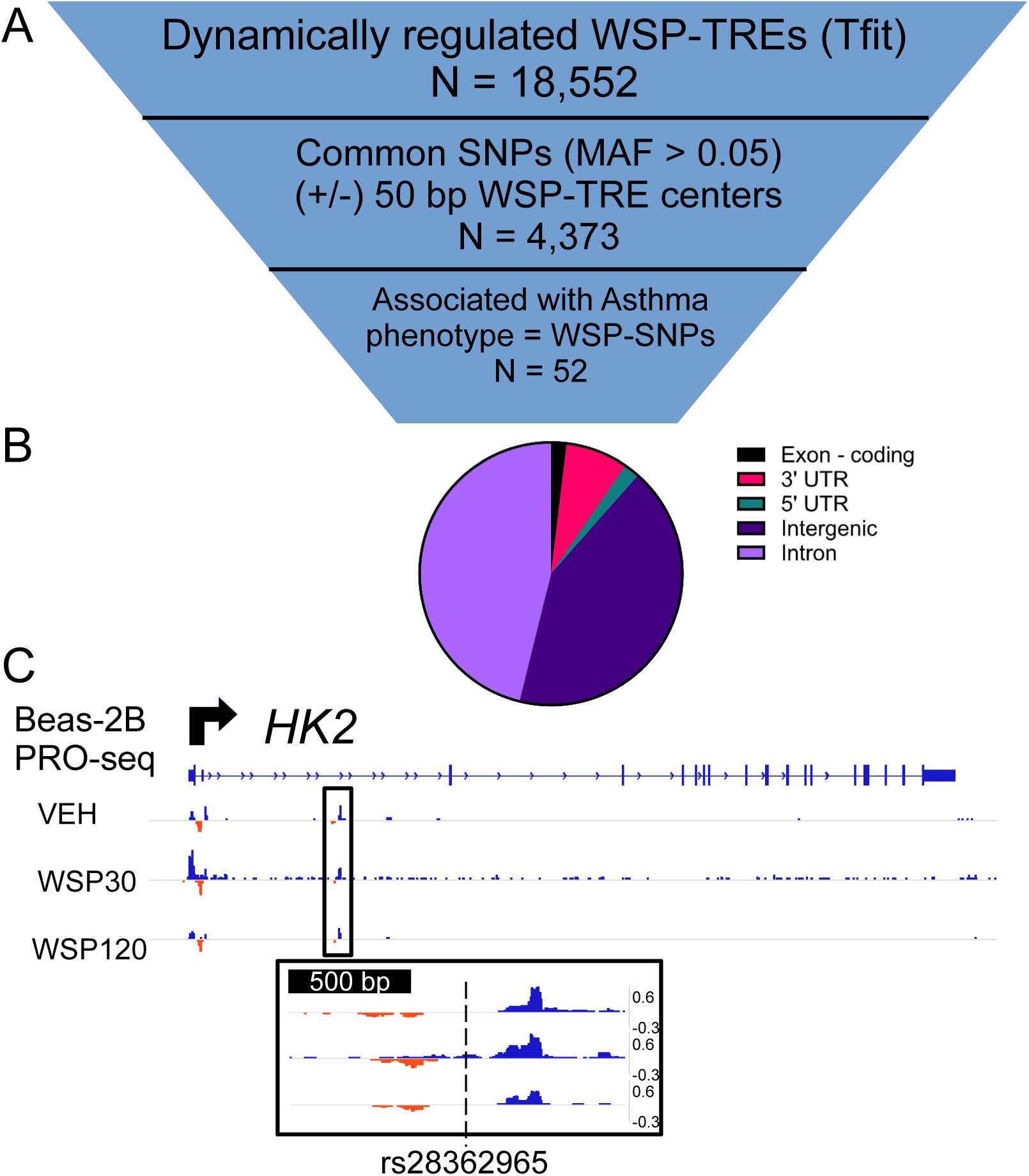
A permutation statistical approach identifies 52 common single nucleotide polymorphisms (SNPs) that reside within transcriptional regulatory elements (TREs) that respond to wood smoke particle (WSP) exposure and associate with asthma diagnosis. A. Schematic of the approach that applies WSP-dependent TREs identified by Precision Run-on sequencing (PRO-seq) in Beas-2B airway epithelial cells to asthma variant discovery. B. Genomic features of the 52 WSP-SNPs. C. Representative example of WSP-SNP which resides within an intronic TRE in the *HK2* gene. PRO-seq data is visualized in the Integrative Genomics Viewer (IGV) genome browser based on counts per million mapped reads (vertical scales). Positive (blue) signal indicates reads annotated to the sense strand while negative (red) signal reflects reads annotated to the antisense strand. The Transcription Start Site (TSS) and direction of transcription are indicated by the arrow at the top of the panel.

**Table 1.**
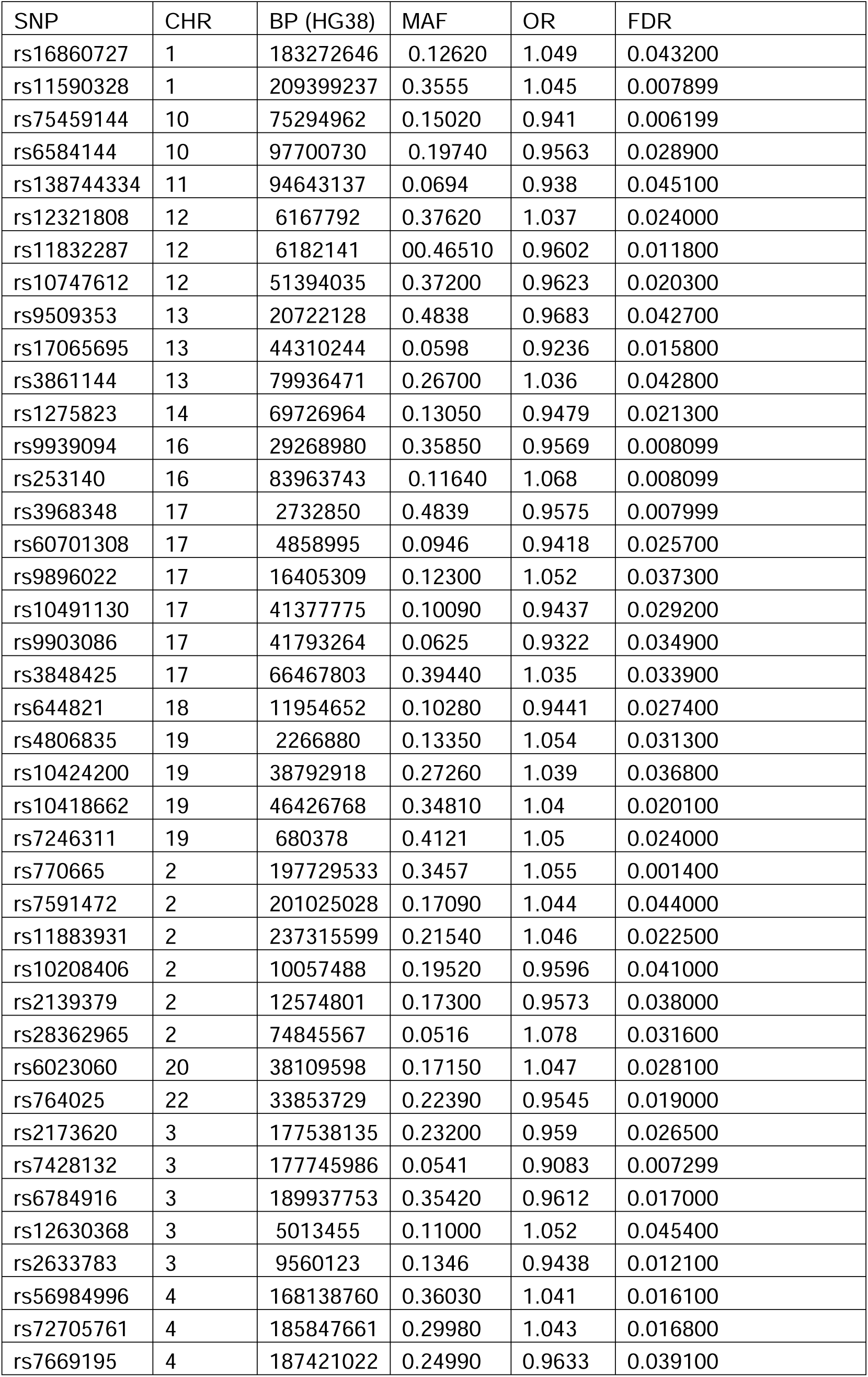
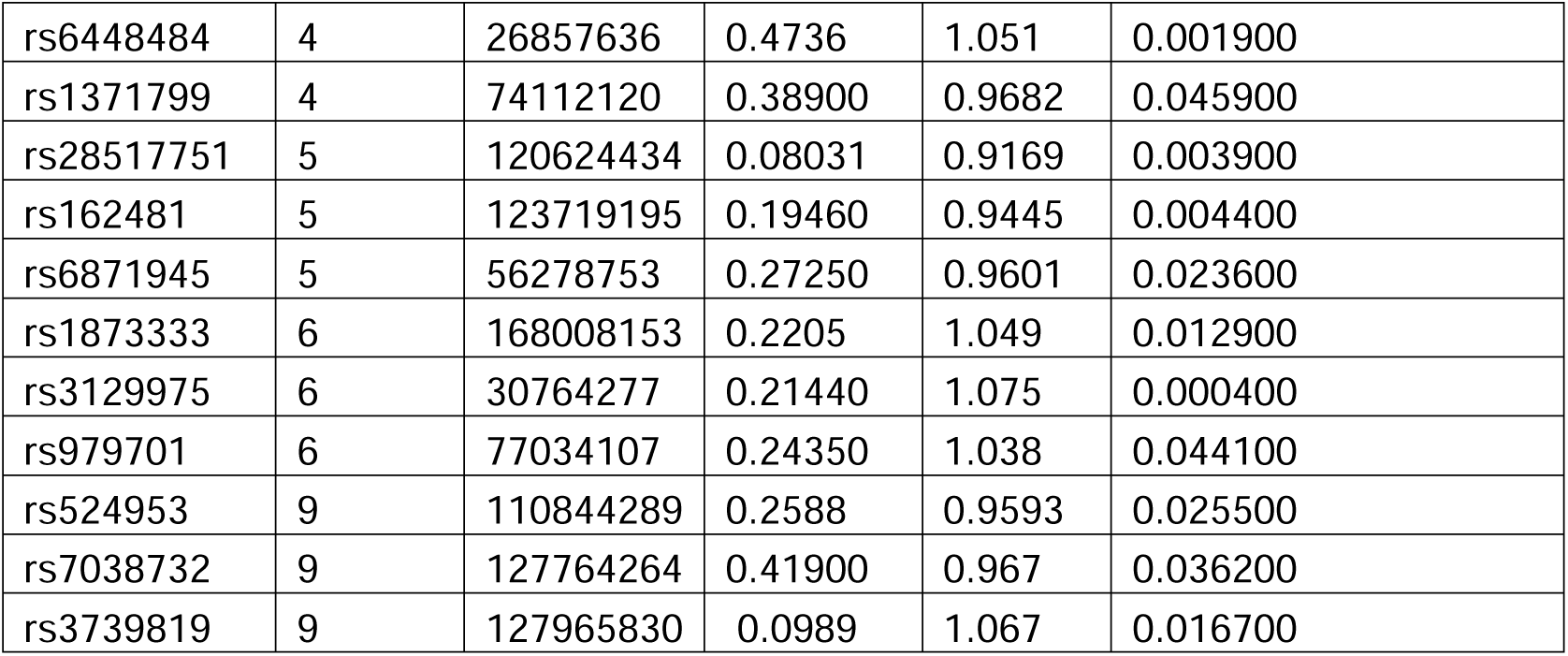
WSP-SNPs.

| SNP | CHR | BP (HG38) | MAF | OR | FDR |
| --- | --- | --- | --- | --- | --- |
| rs16860727 | 1 | 183272646 | 0.12620 | 1.049 | 0.043200 |
| rs11590328 | 1 | 209399237 | 0.3555 | 1.045 | 0.007899 |
| rs75459144 | 10 | 75294962 | 0.15020 | 0.941 | 0.006199 |
| rs6584144 | 10 | 97700730 | 0.19740 | 0.9563 | 0.028900 |
| rs138744334 | 11 | 94643137 | 0.0694 | 0.938 | 0.045100 |
| rs12321808 | 12 | 6167792 | 0.37620 | 1.037 | 0.024000 |
| rs11832287 | 12 | 6182141 | 0.46510 | 0.9602 | 0.011800 |
| rs10747612 | 12 | 51394035 | 0.37200 | 0.9623 | 0.020300 |
| rs9509353 | 13 | 20722128 | 0.4838 | 0.9683 | 0.042700 |
| rs17065695 | 13 | 44310244 | 0.0598 | 0.9236 | 0.015800 |
| rs3861144 | 13 | 79936471 | 0.26700 | 1.036 | 0.042800 |
| rs1275823 | 14 | 69726964 | 0.13050 | 0.9479 | 0.021300 |
| rs9939094 | 16 | 29268980 | 0.35850 | 0.9569 | 0.008099 |
| rs253140 | 16 | 83963743 | 0.11640 | 1.068 | 0.008099 |
| rs3968348 | 17 | 2732850 | 0.4839 | 0.9575 | 0.007999 |
| rs60701308 | 17 | 4858995 | 0.0946 | 0.9418 | 0.025700 |
| rs9896022 | 17 | 16405309 | 0.12300 | 1.052 | 0.037300 |
| rs10491130 | 17 | 41377775 | 0.10090 | 0.9437 | 0.029200 |
| rs9903086 | 17 | 41793264 | 0.0625 | 0.9322 | 0.034900 |
| rs3848425 | 17 | 66467803 | 0.39440 | 1.035 | 0.033900 |
| rs644821 | 18 | 11954652 | 0.10280 | 0.9441 | 0.027400 |
| rs4806835 | 19 | 2266880 | 0.13350 | 1.054 | 0.031300 |
| rs10424200 | 19 | 38792918 | 0.27260 | 1.039 | 0.036800 |
| rs10418662 | 19 | 46426768 | 0.34810 | 1.04 | 0.020100 |
| rs7246311 | 19 | 680378 | 0.4121 | 1.05 | 0.024000 |
| rs770665 | 2 | 197729533 | 0.3457 | 1.055 | 0.001400 |
| rs7591472 | 2 | 201025028 | 0.17090 | 1.044 | 0.044000 |
| rs11883931 | 2 | 237315599 | 0.21540 | 1.046 | 0.022500 |
| rs10208406 | 2 | 10057488 | 0.19520 | 0.9596 | 0.041000 |
| rs2139379 | 2 | 12574801 | 0.17300 | 0.9573 | 0.038000 |
| rs28362965 | 2 | 74845567 | 0.0516 | 1.078 | 0.031600 |
| rs6023060 | 20 | 38109598 | 0.17150 | 1.047 | 0.028100 |
| rs764025 | 22 | 33853729 | 0.22390 | 0.9545 | 0.019000 |
| rs2173620 | 3 | 177538135 | 0.23200 | 0.959 | 0.026500 |
| rs7428132 | 3 | 177745986 | 0.0541 | 0.9083 | 0.007299 |
| rs6784916 | 3 | 189937753 | 0.35420 | 0.9612 | 0.017000 |
| rs12630368 | 3 | 5013455 | 0.11000 | 1.052 | 0.045400 |
| rs2633783 | 3 | 9560123 | 0.1346 | 0.9438 | 0.012100 |
| rs56984996 | 4 | 168138760 | 0.36030 | 1.041 | 0.016100 |
| rs72705761 | 4 | 185847661 | 0.29980 | 1.043 | 0.016800 |
| rs7669195 | 4 | 187421022 | 0.24990 | 0.9633 | 0.039100 |
| rs6448484 | 4 | 26857636 | 0.4736 | 1.051 | 0.001900 |
| rs1371799 | 4 | 74112120 | 0.38900 | 0.9682 | 0.045900 |
| rs28517751 | 5 | 120624434 | 0.08031 | 0.9169 | 0.003900 |
| rs162481 | 5 | 123719195 | 0.19460 | 0.9445 | 0.004400 |
| rs6871945 | 5 | 56278753 | 0.27250 | 0.9601 | 0.023600 |
| rs1873333 | 6 | 168008153 | 0.2205 | 1.049 | 0.012900 |
| rs3129975 | 6 | 30764277 | 0.21440 | 1.075 | 0.000400 |
| rs979701 | 6 | 77034107 | 0.24350 | 1.038 | 0.044100 |
| rs524953 | 9 | 110844289 | 0.2588 | 0.9593 | 0.025500 |
| rs7038732 | 9 | 127764264 | 0.41900 | 0.967 | 0.036200 |
| rs3739819 | 9 | 127965830 | 0.0989 | 1.067 | 0.016700 |

### WSP-SNPs associated with measures of lung function in an independent genetic data set

As an initial validation approach, we analyzed the 52 WSP-SNPs for associations with lung function in previously published GWAS data. We used the Open Targets Genetics platform (27,28) and the Lung Disease Knowledge Portal (29) to navigate GWAS data of lung function measurements, such as forced vital capacity (FVC), forced expiratory volume at 1 second (FEV1), or peak expiratory flow (PEF) (30,31). We identified 6 WSP-SNPs significantly associated (p_adj_ < 0.05) with measures of lung function (Table 2). Three WSP-SNPs (rs3861144, rs3129975 and rs28517751) achieved a statistical threshold for genome-wide association (p < 5×10^-8^). Associations between WSP-SNPs and measurements of lung function, which are objective features of asthma, supported the validity of our discovery data set and motivated further biological study of the 52 WSP-SNPs.

**Table 2.** WSP-SNPs with significant associations in lung function GWAS.

|  |  | Open Targets<br>Genetics | Lung Disease Knowledge<br>Portal |
| --- | --- | --- | --- |
| **SNP** | **Association** | **P value** | **P value** |
| rs3861144 | FEV1 | 6.80E-10 | 8.79E-10 |
| rs28517751 | FEV1/FVC | 4.20E-08 | 4.12E-08 |
| rs3129975 | PEF | 1.30E-26 | 1.74E-25 |
| rs11883931 | FEV1/FVC | 3.90E-05 | 3.36E-05 |
| rs16860727 | FEV1/FVC | 7.60E-06 | 3.80E-06 |
| rs1873333 | PEF | 6.50E-04 | 8.53E-04 |

### Aggregate analysis of the 52 WSP-SNPs suggested regulation of diverse cellular processes

To understand biologic pathways regulated by the WSP-SNPs, we imputed their gene targets using two methods. First, we used expression quantitative trait locus (eQTL) data from the Open Targets Genetics database to assign gene targets to SNPs. This approach was not comprehensive, as multiple WSP-SNPs had no eQTL associations. However, we identified 154 genes associated with at least one WSP-SNP. Functional annotation using DAVID (32) identified no enriched categories for these 154 genes. Second, we applied a permissive approach that identified all genes that were located within 500 kb and shared nascent transcriptional kinetics with the WSP-SNPs’ TRE. This approach identified 376 genes, and one significantly enriched (p_adj_ = 2.1e-7) annotation term – “keratin filament.” A complete list of the putative gene targets from each approach is provided in Supplemental Tables E4 and E5. Based on the limited aggregate clustering, we concluded that the WSP-SNPs regulate diverse mechanisms, and focused analysis was necessary to identify functional SNPs.

### Focused biologic analysis using chromatin accessibility and bioinformatic transcription factor binding predictions identified WSP-SNP candidates for functional analysis

We next applied cellular and molecular features relevant to asthma and wildfire PM2.5 exposures to evaluate regulatory regions containing WSP-SNPs. We found that 23 WSP-SNPs overlapped with chromatin accessibility in live airway epithelial cells obtained from bronchoscopy from three healthy non-smoking adult donors (Figure 2A), supporting the biologic validity of these WSP-SNPs in airway epithelial function. A representative example demonstrates chromatin accessibility in live airway epithelium at the rs9903086 SNP and its neighboring gene *JUP* (Figure 2B). We next scanned genome sequences of TREs containing WSP-SNPs for binding motifs from two classes of transcription factors that are relevant to WSP and PM2.5 exposures – aryl hydrocarbon receptor (AHR) and nuclear factor kappa B (NFkB) (33,34). Using a 500 bp radius to define the regulatory region containing each WSP-SNP, we encountered these motifs at 38 WSP-SNP loci, and 23 WSP-SNP loci contained motifs for both AHR and NFkB factors (Figure 2C), supporting our previous study that demonstrated cross talk between these factors as a mechanism mediating inflammatory responses to WSP exposure (25). The TRE containing rs11883931, which neighbors *LRRFIP1*, is one example that contains both AHR nuclear transporter (ARNT2) and NFkB (REL) transcription factor binding motifs (Figure 2D). These genomic features biologically validated and prioritized a subset of WSP-SNPs for further functional analysis.

**Figure 2.**
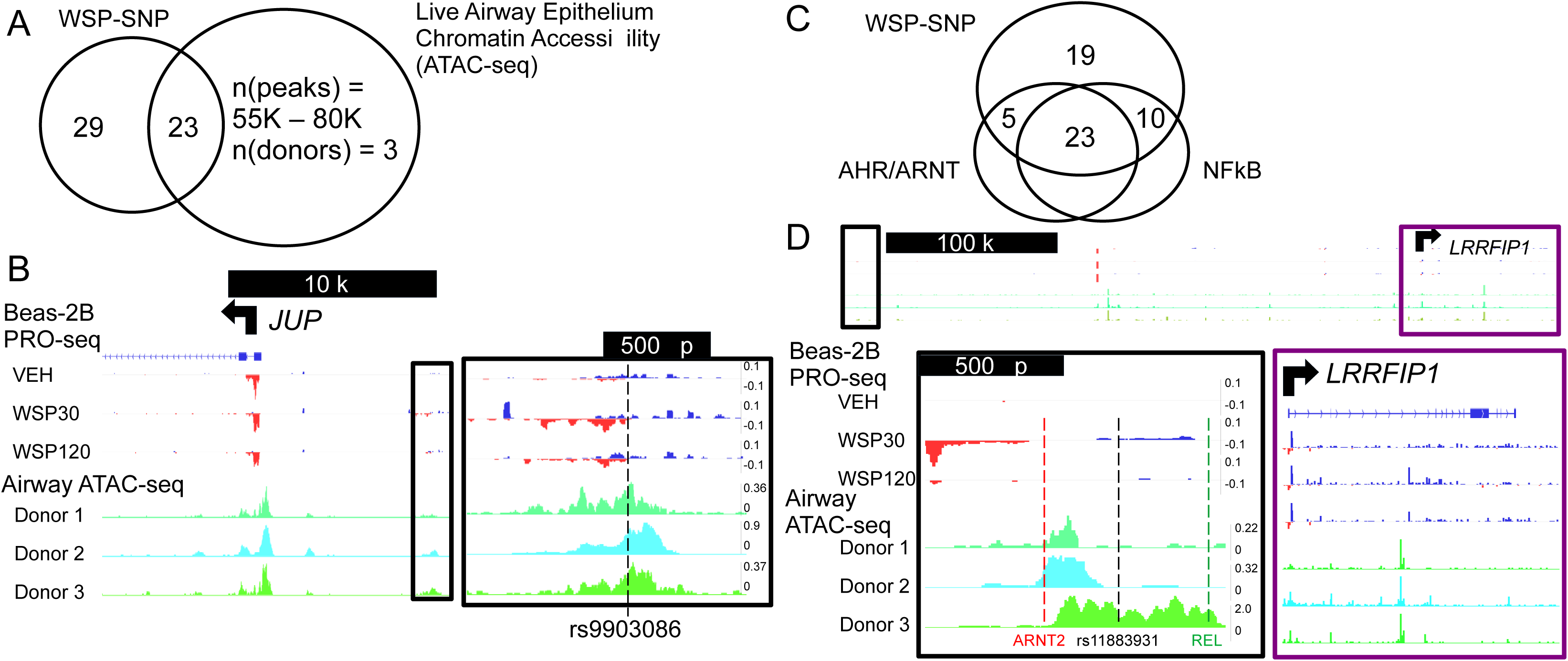
Subsets of the 52 WSP-SNPs exhibit biologic functions associated with asthma. A. Venn diagram demonstrates 23 genomic regions that overlap (> 1 bp), based on genomic coordinates, between WSP-SNPs and chromatin accessibility features in live proximal airway epithelial tissue obtained from healthy non-smoking subjects. B. The representative example rs9903086 WSP-SNP resides in an intergenic region near the *JUP* TSS and overlaps with bidirectional PRO-seq signal in Beas-2B cells and chromatin accessibility signal in live airway epithelium. PRO-seq (blue and red) and ATAC-seq (green and teal) data visualized in the IGV genome browser based on counts per million mapped reads (vertical scales). The TSS is indicated by the arrow at the top of the panel. C. Venn diagram demonstrating sites of genomic overlap between a 500 bp radius surrounding each WSP-SNP and consensus transcription factor binding motifs for aryl hydrocarbon receptor (AHR) and nuclear factor kappa b (NFkB) family transcription factors. Twenty-three WSP-SNP regions contain motifs from both families. D. A representative example at the rs11883931 locus, which contains both ARNT2 (AHR nuclear transporter) and REL (NFkB family) transcription factor binding motifs.

### Functional WSP-SNP genotype changed transcriptional regulatory potential in plasmid reporter systems

We selected 15 prioritized WSP-SNPs to test transcriptional function in plasmid systems (Figure 3A) and cloned 700-800 bp regions surrounding these SNPs into luciferase reporters. Within this reporter set, 5 WSP-SNP constructs exhibited enhancer activity at baseline or in response to WSP exposure, and single base allele change of 2 WSP-SNPs changed the regulatory activity of the entire construct (Figure 3B). Based on this, we defined rs3861144 and rs11832287 as functional genetic variants that alter the transcriptional regulatory activity of their resident TREs based on haplotype.

**Figure 3.**
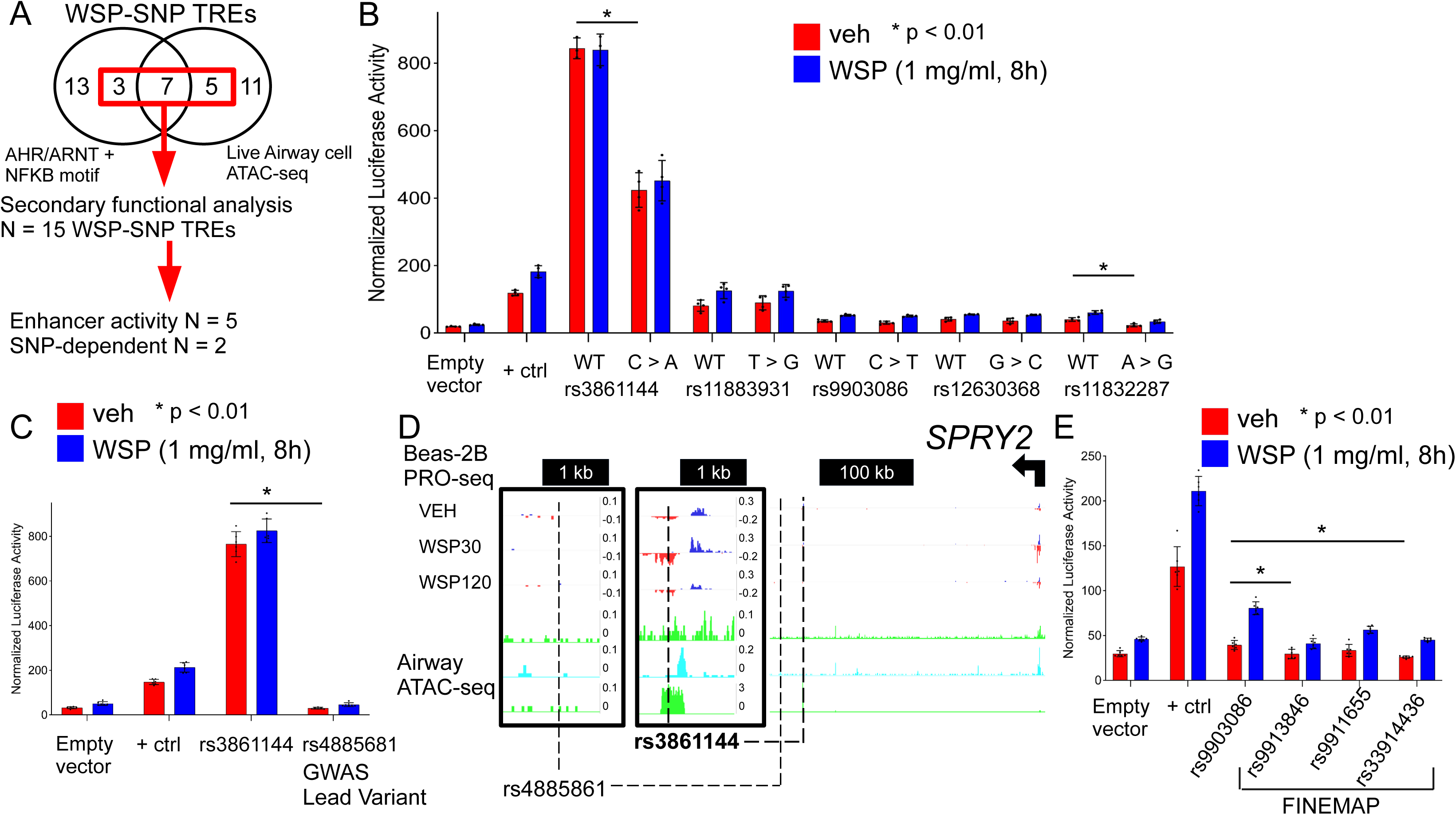
Luciferase activity in plasmid reporter systems demonstrates that WSP-SNPs reside in active TREs in comparison with local lead variants predicted by GWAS. A. Schematic for selection of WSP-SNP candidates prioritized by biologic variables of live airway chromatin accessibility and colocalization with both AHR and NFkB family transcription factor binding motifs. B. Normalized luciferase activity of plasmid reporters transfected into Beas-2B cells and treated with vehicle (veh) or WSP (1 mg/ml) for 8 hrs. A 700 – 800 bp genomic region surrounding each WSP-SNP was cloned into the plasmid system and mutated for allele change, with “WT” indicating the allele native to Beas-2B cells and genotype letter change indicating the mutated reporter. Bars indicate mean reporter activity (+/-standard deviation, n = 4) relative to an internal control Renilla luciferase that was co-transfected into cells; * p < 0.01. C. Normalized luciferase activity of plasmid reporters containing the WSP-SNP rs3861144 and rs4885681, the lead variant at this locus prioritized by GWAS statistics. A 350 – 400 bp radius genomic region surrounding each SNP was cloned into the plasmid system, and luciferase activity was measured after transfection into Beas-2B cells and treatment with vehicle (veh) or WSP (1 mg/ml) for 8 hrs. Data are presented as described for (B). D. Beas-2B PRO-seq and live airway epithelium ATAC-seq signal at the WSP-SNP rs3861144 and rs4885681 loci, located approximately 380 kb 3’ to the *SPRY2* gene. E. Normalized luciferase activity of plasmid reporters containing a 350-400 bp radius around the WSP-SNP rs9903086 and three SNPs (rs9913846, rs991655 and rs39914436) prioritized by statistical fine mapping (FINEMAP (35)) for association with Forced Vital Capacity using summary statistics from an independent data set (30). Bars indicate mean reporter activity (+/-standard deviation, n = 4) relative to Renilla; * p < 0.01.

### WSP-SNPs identified functional variants at statistically prioritized GWAS loci

The functional genetic variants that we identified were not previously reported but had statistical associations with lung function, such as rs3861144 with FEV1 (Table 2). We next identified the leading statistical candidate SNPs that colocalize with WSP-SNPs to compare regulatory functions using the luciferase reporter system. We performed Bayesian fine mapping (35) for the continuous FVC trait (36) at the rs3861144 and rs9903086 loci. No SNPs were identified in the credible set at rs3861144, but rs4885681 was identified as the local lead variant based on the lowest p value for association (p = 1.3×10^-11^). The rs3861144 plasmid reporter had greater transcriptional regulatory activity in the luciferase assay when compared with a plasmid reporter similarly constructed for rs4885681, which resides in a non-annotated and non-functional genomic region (Figure 3C-D). We selected three SNPs within the credible set (posterior probability > 0.95) of association with FVC at the rs9903086 locus – rs9913846, rs9911655, and rs33914436 (Supplemental Table E6). Again, the transcriptional regulatory activity of the rs9903086-containing plasmid reporter at baseline and in response to WSP exposure was greater than reporters containing these local fine-mapped SNPs (Figure 3E), which reside in genomic regions without functional annotations (Supplemental Figure E1). These results demonstrate that within a genomic locus associated with lung function, the genetic variants identified by our approach have greater transcriptional regulatory function than the lead variants reported by prior GWAS.

### Shared nascent transcriptional kinetics between gene and TRE identified SPRY2 as the target of the rs3861144 WSP-SNP

We applied PRO-seq data, three-dimensional chromatin analysis, and genome editing to propose and confirm the gene target of one selected WSP-SNP, rs3861144. Prior studies have connected TREs with target genes based on shared activity and genomic proximity (37). Using this concept, the regulatory element containing rs3861144, a WSP-SNP independently associated with lung function with allele-dependent transcriptional regulatory activity, regulates *SPRY2* in response to WSP exposure (Figure 4A). Three-dimensional chromatin analysis using micrococcal nuclease chromosome conformation capture (Micro-C) in unstimulated Beas-2B cells (38) demonstrated that rs3861144 and *SPRY2* reside in the same topologically associated domain and may interact indirectly (Supplemental Figure E2). To definitively link the TRE harboring rs3861144 with control of *SPRY2*, we deleted the rs3861144 TRE using CRISPR-Cas9 and analyzed *SPRY2* expression using qPCR in Beas-2B cells. PCR amplification of genomic DNA from cells transfected with negative control or rs3861144-targeting guide RNA demonstrated efficient deletion of this TRE (Figure 4B). Cells with rs3861144 TRE deletion showed a reduction in *SPRY2* expression response to WSP (Figure 4C). These results connected the rs3861144 TRE with *SPRY2* expression in response to WSP and demonstrated the facility of functional SNPs to connect exposure, gene mechanism, and phenotype.

**Figure 4.**
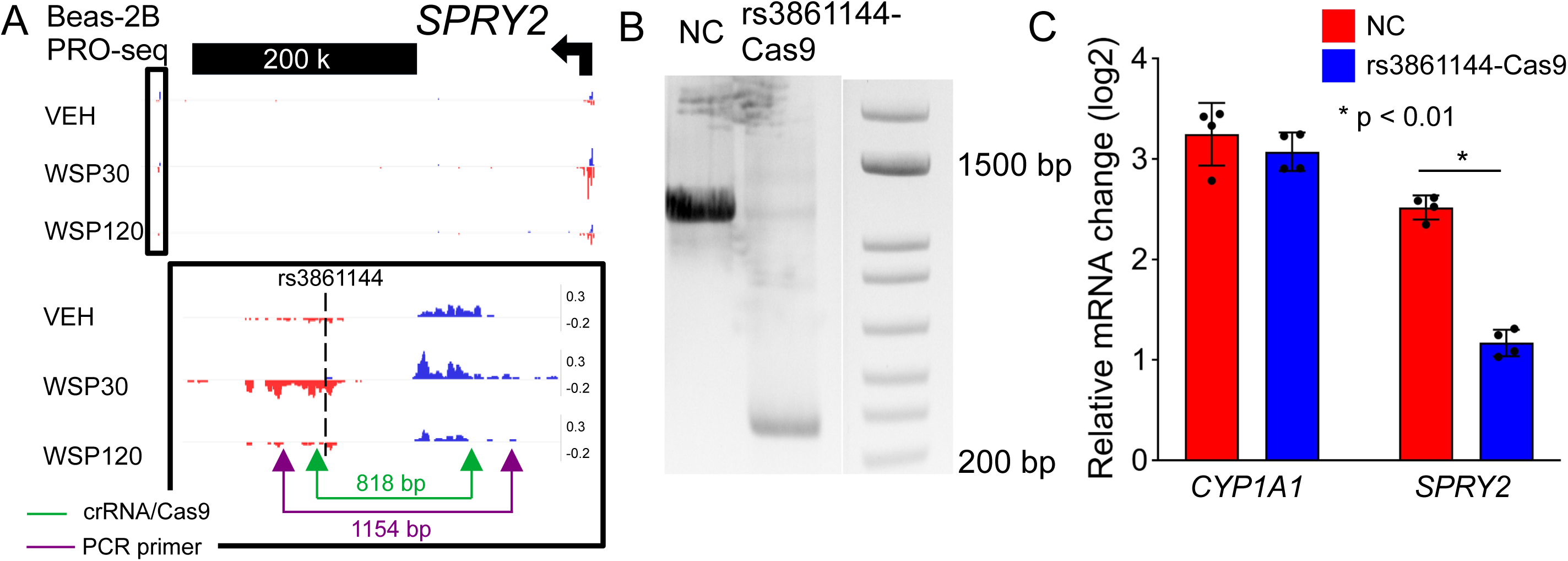
The rs3861144 functional variant resides in a TRE that regulates *SPRY2* in response to WSP exposure in Beas-2B cells. A. Beas-2B PRO-seq signal visualized in IGV at the rs3861144 and *SPRY2* gene locus demonstrates shared transcriptional kinetics between these regions at 30 and 120 minutes of WSP exposure. Below – green arrows indicate two target guide RNA (gRNA) sites designed for Cas9 genome editing of the rs3861144 TRE; purple arrows indicate target sites for polymerase chain reaction (PCR) primers used to amplify the region following Cas9 editing and confirm genomic deletion. B. Beas-2B cells were transfected with two on-target Cas9-gRNA RNP complexes indicated by green arrow sites in the rs3861144 TRE in (A) (rs3861144-Cas9) or negative control Cas9-gRNA RNP (NC). Gel electrophoresis confirmed genomic deletion at the rs3861144 target region in cells transfected with targeting gRNAs. C. CRISPR-Cas9 transfected cells were exposed to WSP at 1 mg/ml (0.26 mg/cm^2^) for 2 hours. Gene expression of the canonical WSP target *CYP1A1* and *SPRY2* were measured using quantitative PCR (qPCR). Bars represent mean normalized C_T_ values on a log_2_ scale (+/-standard deviation, n = 4) relative to vehicle-treated controls; * p < 0.01.

### Cytokine kinetics are regulated by SPRY2 knockdown in Beas-2B cells exposed to WSP

We next studied how *SPRY2* knockdown interacts with WSP exposure by applying short-interfering RNA targeted to *SPRY2* (si*SPRY2*) in Beas-2B cells and WSP exposure at multiple time points. We first performed RNA sequencing in si*SPRY2* and control cells exposed to WSP for 2 and 4 hours. A principal component plot of sample clustering is presented in Supplemental Figure E4. Within gene expression differences between WSP-exposed si*SPRY2* and control cells at 4 hours of exposure, we observed 180 differentially expressed genes enriched in si*SPRY2* cells (p_adj_ < 0.05, log_2_FoldChange > 0.5), and using DAVID (32) for functional annotation, we identified “Growth Factor” (FDR = 0.028) and “TNF signaling” (FDR = 0.023) terms enriched in this gene set. Supplemental Table E7 contains a complete list of significant functional annotation terms (FDR < 0.05). These results were supported by changes in ERK1/2 phosphorylation kinetics in si*SPRY2* cells compared with controls under acute WSP exposure (Supplemental figure E5). Next, independent comparisons in si*SPRY2* and control cells demonstrated 103 genes with increased expression between 2- and 4-hour WSP exposures in control cells and 180 genes in si*SPRY2* cells (Figure 5B-C). Functional annotation terms from these gene sets had notable differences in cytokine enrichment, which suggested an interaction between si*SPRY2* and cytokine gene expression kinetics (Table 3, Supplemental Tables E8-E9). However, when we tested Interleukin-8 as a representative cytokine in si*SPRY2* and control cells exposed to WSP for 24 hours, we observed reduced *CXCL8* gene expression and reduced IL8 cytokine secretion by Enzyme Linked Immunosorbent Assay in WSP-exposed si*SPRY2* cells (Figure 5D, Supplemental Figure E6). This change was consistent with a reduction in ERK1/2 phosphorylation in si*SPRY2* cells at later time points (Figure 5E), and no significant cytotoxicity was observed at this time point (Supplemental Figure E7). These results indicated that *SPRY2* knockdown exhibits variable interactions with cytokine gene expression in Beas-2B cells exposed to WSP, and sustained WSP exposure results in reduced IL8 secretion and ERK1/2 phosphorylation in cells with *SPRY2* knockdown.

**Figure 5.**
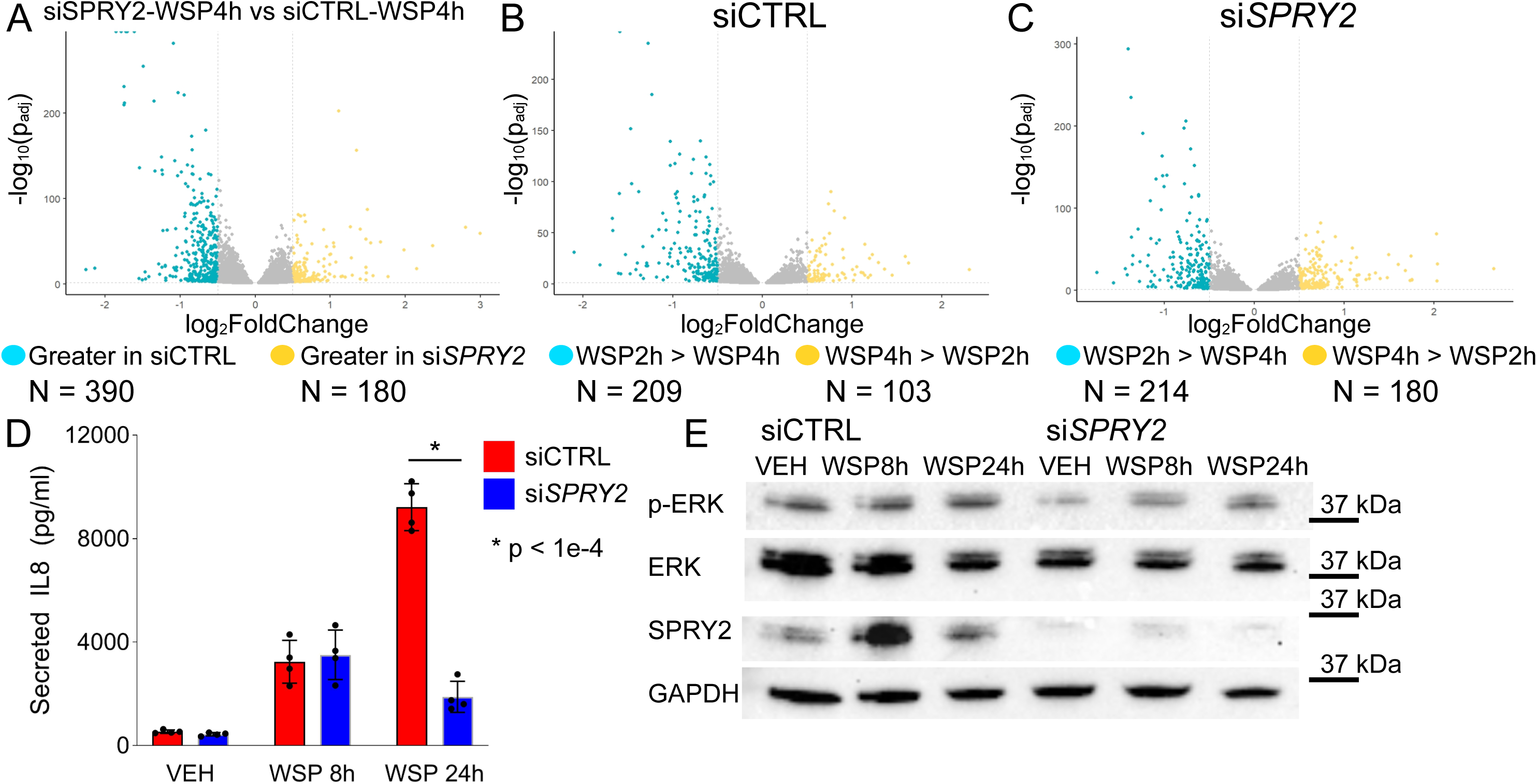
*SPRY2* knockdown by siRNA results in acute and latent responses to WSP exposure. A. Volcano plot from RNA-sequencing data demonstrates differentially expressed genes between WSP-exposed (4 hours) non-targeting control (siCTRL) or *SPRY2*-targeting siRNA (si*SPRY2*) transfected Beas-2B cells. Each point represents a single gene, with the x-axis showing log_2_ fold change and the y-axis showing the log transformed p_adj_ statistic. Dashed lines indicate log_2_ fold change of 0.5, which was the threshold used for this analysis. Number of differentially expressed genes (p_adj_ < 0.05 and log_2_ fold change > 0.5) are listed below the plot. B. Volcano plot demonstrates differentially expressed genes between 2- and 4-hour WSP exposure in siCTRL cells. C. Volcano plot demonstrates differentially expressed genes between 2- and 4-hour WSP exposure in si*SPRY2* cells. D. ELISA analysis of IL-8 concentration in the supernatant of siCTRL and si*SPRY2* Beas-2B cells exposed to WSP (1 mg/ml of cell culture media or 0.26 mg/cm^2^ cell culture surface) at latent time points. Bars indicate mean IL-8 concentration (+/-SD); n = 4/group, *p < 1×10^-4^ for indicated comparisons. E. Western blot demonstrates knockdown in *SPRY2* expression with *SPRY2*-siRNA transfection (si*SPRY2*) is associated with reduced phosphorylated ERK1/2 in vehicle- and WSP-exposed (24 hour) Beas-2B cells compared with control-transfected cells (siCTRL). GAPDH served as the loading control.

**Table 3.**
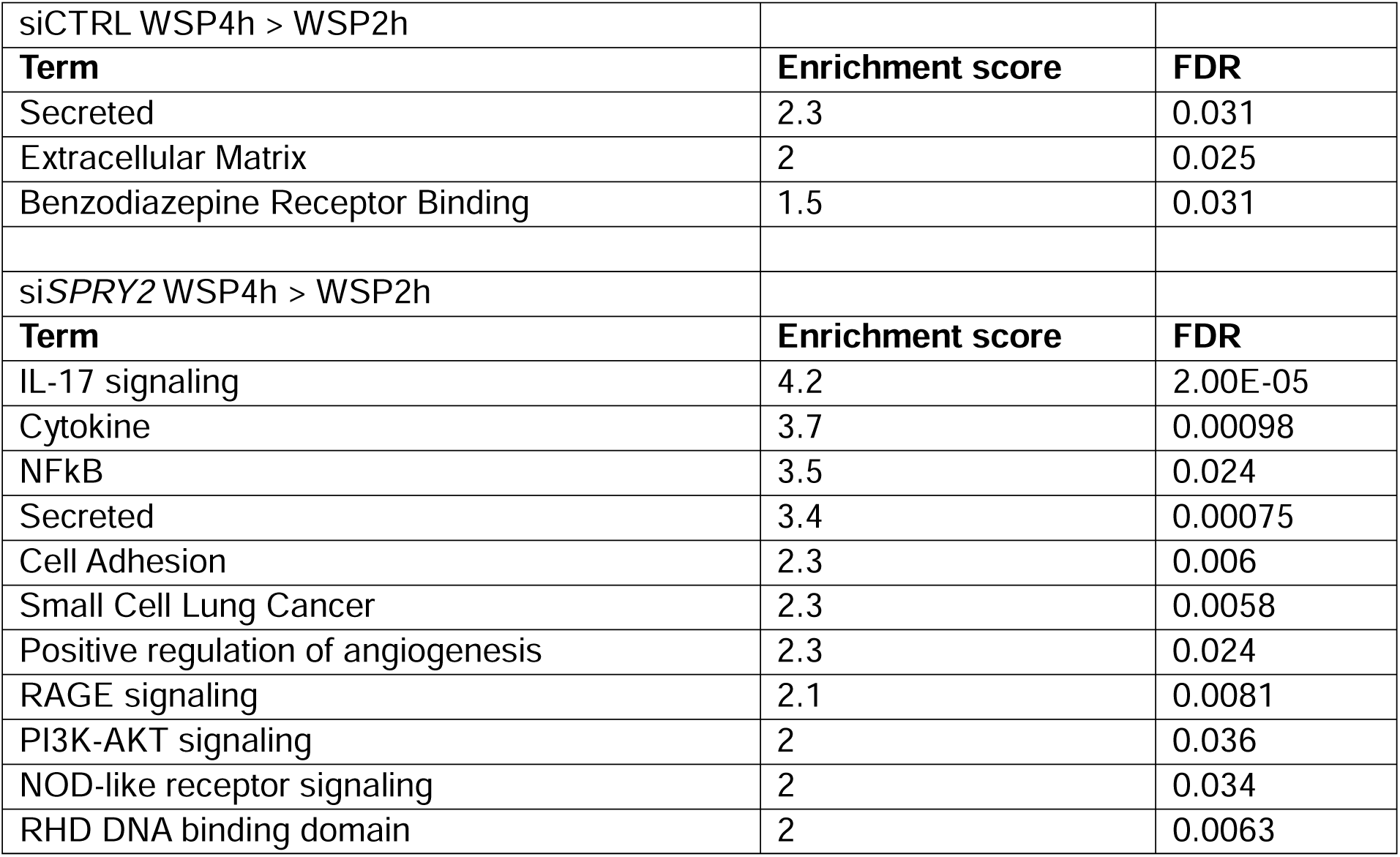
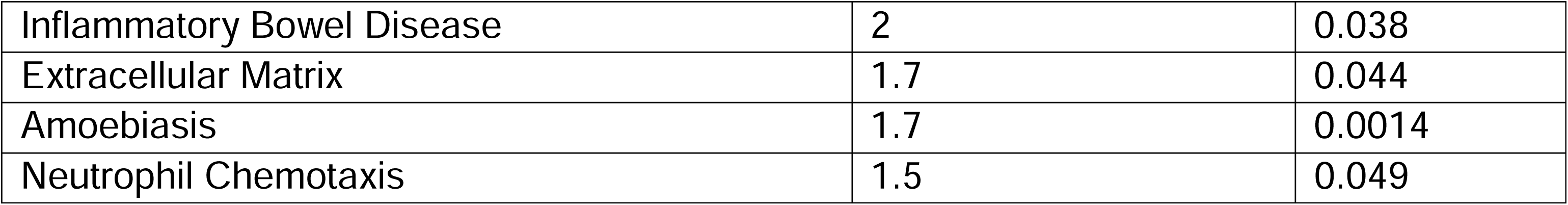
Functional annotation categories for genes upregulated between 2 and 4 hours of WSP exposure in control (siCTRL) and *SPRY2* knockdown (si*SPRY2*) cells.

| siCTRL WSP4h > WSP2h |  |  |
| --- | --- | --- |
| **Term** | **Enrichment score** | **FDR** |
| Secreted | 2.3 | 0.031 |
| Extracellular Matrix | 2 | 0.025 |
| Benzodiazepine Receptor Binding | 1.5 | 0.031 |
| siSPRY2 WSP4h > WSP2h |  |  |
| **Term** | **Enrichment score** | **FDR** |
| IL-17 signaling | 4.2 | 2.00E-05 |
| Cytokine | 3.7 | 0.00098 |
| NFkB | 3.5 | 0.024 |
| Secreted | 3.4 | 0.00075 |
| Cell Adhesion | 2.3 | 0.006 |
| Small Cell Lung Cancer | 2.3 | 0.0058 |
| Positive regulation of angiogenesis | 2.3 | 0.024 |
| RAGE signaling | 2.1 | 0.0081 |
| PI3K-AKT signaling | 2 | 0.036 |
| NOD-like receptor signaling | 2 | 0.034 |
| RHD DNA binding domain | 2 | 0.0063 |
| Inflammatory Bowel Disease | 2 | 0.038 |
| Extracellular Matrix | 1.7 | 0.044 |
| Amoebiasis | 1.7 | 0.0014 |
| Neutrophil Chemotaxis | 1.5 | 0.049 |

### SPRY2 knockdown resulted in impaired mechanical scratch repair in Beas-2B cells

Given the association between *SPRY2* knockdown and reduced IL8 responses to WSP exposure and prior reports associating *SPRY2* with growth factors and cell proliferation (39–41), we next studied the interaction between *SPRY2* knockdown and epithelial monolayer injury by mechanical scratch. In the absence of WSP exposure, we measured the closure of a mechanical scratch defect in submerged culture of si*SPRY2* and control Beas-2B cells. Scratch area in si*SPRY2* cells separated from control cells at later time points, with the largest difference at 16 hours following injury (Figure 6A), consistent with later time points that demonstrated reduced IL8 secretion following WSP exposure. Representative images from one experiment are presented in Figure 6B. We again observed reduced ERK1/2 phosphorylation in unstimulated proliferating si*SPRY2* cells compared with controls (Figure 6C). These results established the functional significance of *SPRY2* knockdown and rs3861144 in airway epithelial cell physiology.

**Figure 6.**
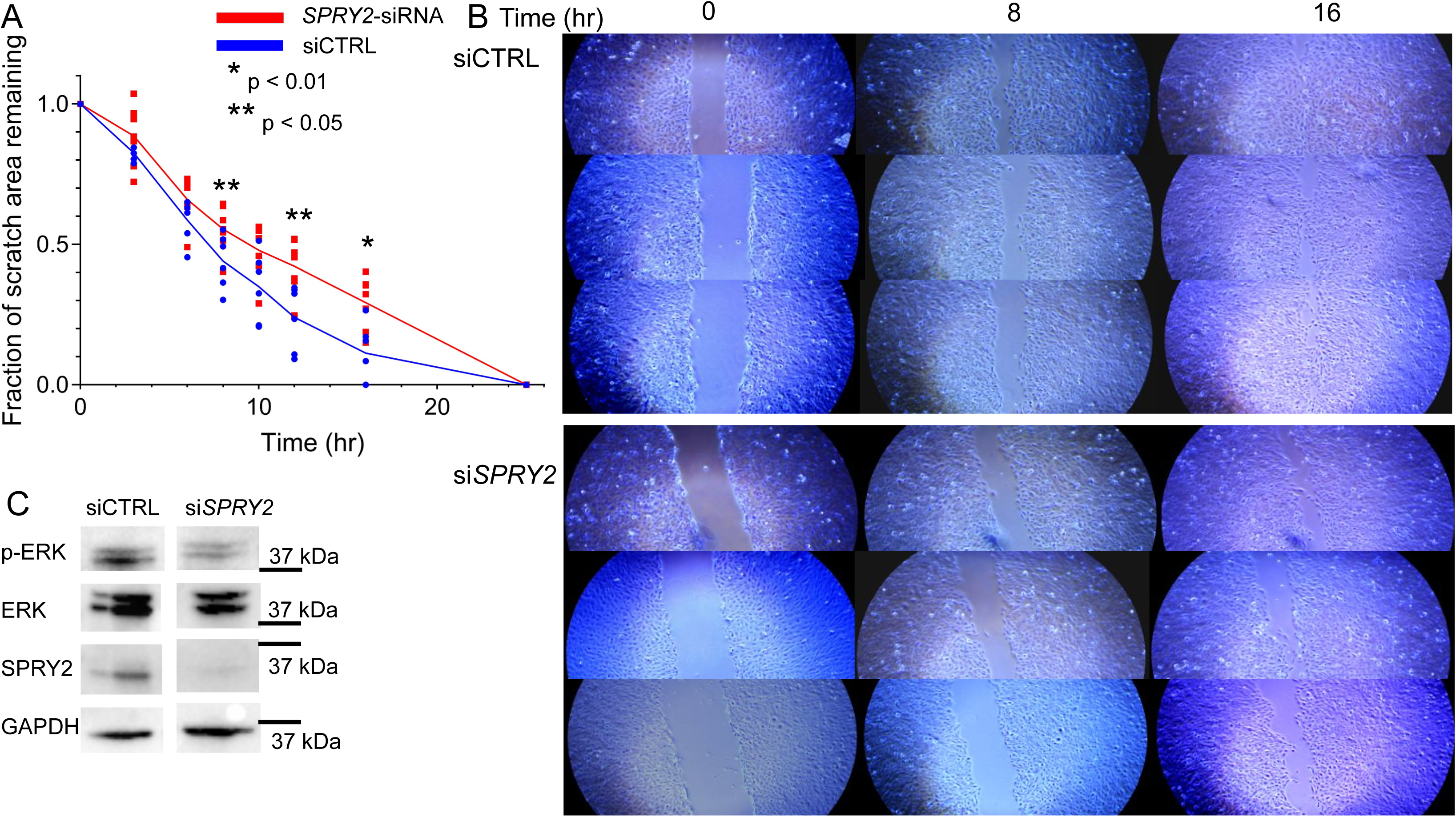
*SPRY2* knockdown by siRNA delays scratch repair in a Beas-2B monolayer. A. Scratch assay in siCTRL or si*SPRY2* Beas-2B cells demonstrated a larger remaining defect at 8, 12, and 16 hours in cells with *SPRY2* knockdown. Solid line depicts the interpolated slope between the means of measurements at 2, 6, 8, 10, 12, 16 and 25 hours. Individual measurements are plotted as blue (non-targeting control “siCTRL”) or red (*SPRY2*-siRNA) points (n=6-7/condition). * p < 0.01, ** p < 0.05. B. Representative images from three wells of siCTRL and si*SPRY2* scratch assay at three time points. C. Western blot demonstrates knockdown in *SPRY2* expression in si*SPRY2* cells is associated with a reduction in phosphorylated ERK1/2 signal in siCTRL cells. GAPDH served as the loading control.

### SPRY2 is regulated in primary human airway cells

To determine whether *SPRY2* is regulated in primary human airway cells, we next examined responses in relevant primary human airway models. We demonstrated that *SPRY2* expression increases in primary SAECs cultured and exposed to WSP in submerged/basal conditions (Figure 7A). The gene expression kinetics of *SPRY2* were similar to kinetics in Beas-2B cells, peaking at 2 hours and decreasing by 4 hours of WSP exposure. We also demonstrated that *SPRY2* expression was regulated in primary human airway smooth muscle (HASM) cells in response to TNF-alpha, a secondary mediator of WSP released by airway epithelial and innate immune cells, and primary HASM cells from one asthma donor exhibited reduced *SPRY2* expression (Figure 7B). Finally, we performed *SPRY2* knockdown using CRISPR-Cas9 in SAECs from one donor (Sm36) and demonstrated a reduced ratio of phosphorylated to dephosphorylated ERK1/2 in *SPRY2*-knockdown basal SAECs. (Supplemental Figure E9). Therefore, *SPRY2* is regulated in primary human airway cells and controls MAPK signaling by regulating ERK1/2 expression and phosphorylation.

**Figure 7.**
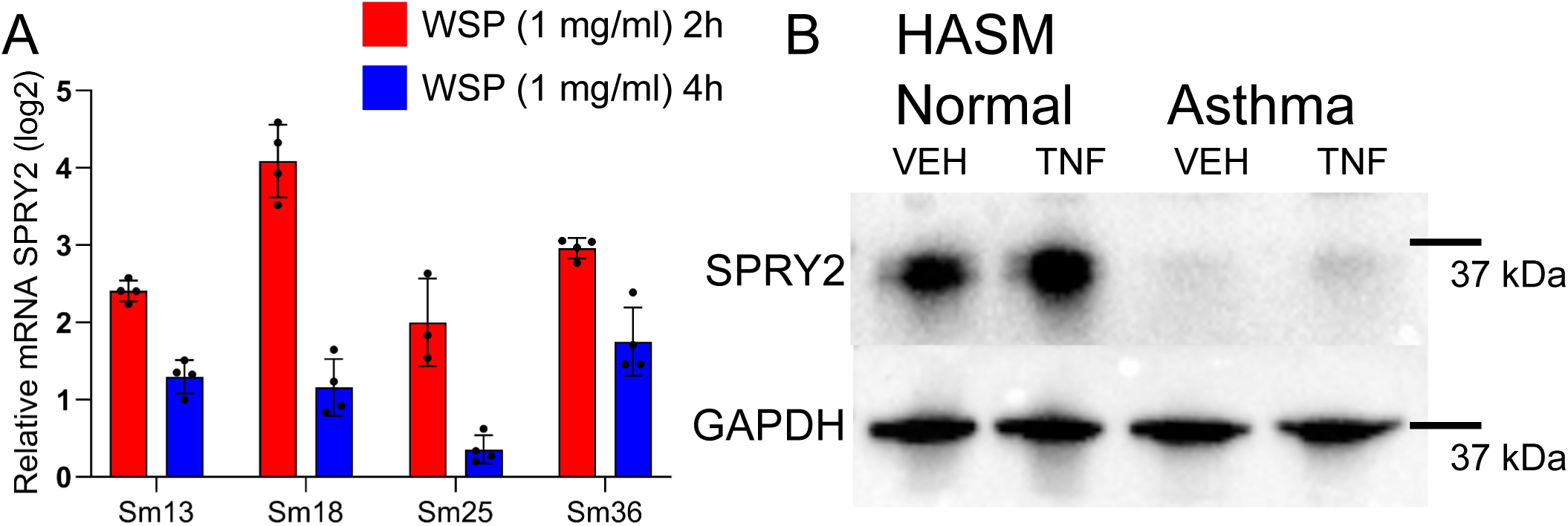
*SPRY2* is expressed and regulated in primary human airway cells. A. Primary small airway epithelial cells obtained from four healthy, non-smoking donors were exposed to WSP (1 mg/ml, 0.26 mg/cm^2^) in submerged culture for 2 or 4 hours. Gene expression for *SPRY2* was measured using qPCR. Bars represent mean normalized C_T_ values on a log_2_ scale (+/-standard deviation, n = 4) relative to vehicle-treated controls. B. *SPRY2* expression in normal HASM cells is increased by TNF-alpha stimulation but reduced in cells from an asthmatic donor. GAPDH was the loading control.

## Discussion

Wildfire related PM2.5 air pollution is increasing and has a direct impact on asthma morbidity (10,42,43). However, the genetic and molecular mediators of these malign effects are poorly understood. To address these gaps, we developed a genetic approach to discover novel relationships between asthma, SNPs, and gene expression responses to WSP exposure in an airway epithelial cell culture model. Through this approach, we identified 52 common variant SNPs within WSP-responsive TREs that were nominally associated with an asthma diagnosis in the GERA cohort. We prioritized SNPs based on primary human airway chromatin accessibility profiles and bioinformatic predictions of transcription factor binding, and we identified two functional asthma variants, rs3861144 and rs11832287. Transcriptional regulatory activity of the regions harboring rs3861144 and rs11832287 was controlled by haplotype, and based on congruent TRE and gene transcriptional responses, we identified the putative gene targets of rs3861144 and rs11832287 as *SPRY2* and *CD9*. We confirmed the regulatory association between rs3861144 and *SPRY2* using a CRISPR-based deletion strategy, and we demonstrated that *SPRY2* is a plausible candidate for asthma pathobiology through cellular functions that include regulating ERK1/2 phosphorylation, cytokine secretion, and monolayer scratch repair. Our findings establish a powerful pipeline for genetic discovery of associations between asthma and woodsmoke and highlight a novel link between *SPRY2* regulation by WSP and risk of an asthma diagnosis.

Our pipeline for genetic discovery builds on genome-wide association studies by re-analyzing existing data to narrow the focus to functional variants, which can be more directly connected with gene targets than statistically prioritized results. We first used PRO-seq-annotated WSP responses to pre-select SNPs to test for asthma association that reside in the functional centers of TREs, which are enriched for transcriptional machinery (44,45) and regulatory SNPs (46,47). This constraint enriched our discovery for functional SNPs and reduced the genomic sequences for discovery to a total of ∼1 Mb. The reduced number of total SNPs tested allowed us to apply permissive permutation-based statistics and identify variants that would not meet genome-wide statistical thresholds. Previous studies that have applied genomic annotations to prioritize functional variants from GWAS results (48–51) are limited in only analyzing variants that meet these rigorous statistical thresholds and may miss functional variants with weaker statistical associations. Second, we pre-selected SNPs that reside in WSP-responsive TREs to enrich genetic discovery for SNPs whose transcriptional regulatory activity depends on WSP exposure in cell culture models. We further prioritized SNPs based on proximity to regulatory motifs that are well-established to mediate responses to combusted particles, such as wood smoke, *in vivo*. While this method does not directly integrate the complexity of environmental exposures with the genetic risk of asthma, it does provide insight into asthma risk in the context of genomic responses to relevant exposures.

The use of PRO-seq annotations in our pipeline facilitated connecting functional variants with target genes that respond to pertinent exposures based on transcriptional kinetics. The location of most genetic risk variants in non-coding and non-annotated genomic regions (52) presents a general challenge for defining relevant target genes and associated mechanisms from GWAS results. Expression quantitative trait locus (eQTL) analysis can address this problem (53) but requires knowledge of the functional cell type and access to large tissue gene expression data sets. Direct exposure measurements in relationship to gene expression are also challenging for eQTL studies, and unaccounted variable exposures can confound eQTL results. We connected functional genetic variants with gene targets by applying PRO-seq data from our cell exposure model to an established paradigm for regulatory element-gene association (37). The advantages of our pipeline relative to other methods led to the discovery of rs3861144 and *SPRY2* as novel mechanisms that connect WSP with asthma. Future studies that integrate our method with existing tools may facilitate additional discovery of variants and target genes that influence gene-by-environment risk in asthma.

We demonstrated the utility of our genetic approach by prioritizing rs3861144 as the functional variant within its locus and connecting the disease variant with a reduced *SPRY2* gene expression response to WSP exposure. *SPRY2* is a kinase modulator of the mitogen-activated protein kinase (MAPK) pathway that plays diverse cellular roles in regulating cell growth, survival, and proliferation (54–56). Prior reports have connected *SPRY2* gene regulation with functional consequences (41,57). Previous studies have also reported that genetic signals in the *SPRY2* locus control lung function. Multiple linked SNPs, including rs3861144, were statistically associated (p < 5×10^-9^) with measurements of lung function, such as FEV1 (36). However, this study prioritized rs4885681 as the lead SNP based on the strongest association statistics and connected rs4885681 with the LINC00382 long non-coding RNA, based on its genomic location. We demonstrated that rs4885681 did not exhibit features of a functional genetic variant, such as genomic annotations or transcriptional regulatory function in luciferase reporters. Therefore, the mechanism derived from rs4885681 was unlikely to explain the connection between genetic signals at this locus and lung function. In contrast, we prioritized rs3861144 as a functional variant at this locus, and using nascent transcript annotations and genome editing, we identified *SPRY2* as its gene target and the plausible mechanism that connected the genetic signals in this locus with lung function. These data demonstrate that defining the functional variants within a statistically associated genetic locus provides additional utility to GWAS in connecting gene mechanisms with phenotypes.

We also demonstrated changes in airway epithelial cell physiology associated with *SPRY2* knockdown that suggest rs3861144 relates to asthma mechanisms. In our model system, sustained WSP exposure resulted in increased *SPRY2* protein expression at latent time points (8-24 hours), and at these latent time points, *SPRY2* knockdown resulted in reduced IL8 secretion, reduced ERK1/2 phosphorylation, and an impaired ability to close a mechanical scratch in submerged cell culture. These results were consistent with prior reports in airway epithelial cells (58) and rhabdomyosarcoma cell lines (59) and suggest that rs3861144 and *SPRY2* have functional roles in the airway epithelium that relate to asthma pathobiology. We also observed a distinct context associated with acute WSP exposure, where *SPRY2* knockdown increased ERK1/2 phosphorylation and cytokine gene expression kinetics between 2-6 hours of WSP exposure. Some of these changes, such as increased *CXCL8* gene expression in *SPRY2* knockdown cells, were not recapitulated in functional signals, such as IL8 secretion, suggesting that the latent program was more relevant to *SPRY2* function. However, our observations were consistent with prior studies that have reported that *SPRY2* function is highly dependent on cell context, such as growth factor presence (41), and further study of the acute context is necessary to resolve the complex roles of *SPRY2* in the airway epithelium.

Our study had several limitations. The positive predictive value of our primary discovery in relationship to our secondary analyses was modest. Only 6/52 SNPs had independent associations with lung function and 2/52 SNPs were rigorously defined as functional variants. Low thresholds and limitations in accurately defining TREs bioinformatically and coding the asthma diagnosis may have contributed to these results, as well as our application of lung function genetics as a surrogate validation tool for our primary findings. We also performed our initial genetic analyses in mostly non-Hispanic white cohorts and used a single reference for linkage disequilibrium analysis, limiting the generalizability of the results and increasing potential bias. Future validation in an independent multi-ancestry asthma cohort with a whole genome sequencing data set will improve the rigor and confidence of the results.

Our WSP and cell culture exposure system also had limitations. We used aged, desiccated, and UV-exposed WSP from a single combustion source, and the WSP dose that we selected (1 mg/ml or 0.26 mg/cm^2^) lacks a specific *in-vivo* correlation. We cited prior studies of *in-vivo* exposure (22), comparison with established toxicological doses of TCDD (60,61), and the induction of relevant pathways, such as aryl hydrocarbon receptor and inflammasome signaling (62–64), as evidence to support our use of this dose. However, this concentration of WSP promotes a bias towards cell stress pathways and away from potentially important metabolic responses, which have also been associated with asthma (65), and limits in the interpretation of our results overall. Inconsistencies between the Beas-2B immortalized airway epithelial cell line and normal human airway epithelia, such as supraphysiologic concentrations of growth factors (66), lack of active metabolic pathways (67), cilia, and mucus functions (68) also resulted in a bias towards transcriptional programs involved in cell proliferation in our model exposure system. Lacking from our model were important airway epithelial programs, such as mucus dysregulation, metabolism, and cell junction integrity, which also contribute to asthma development (69–72). The acute 30- and 120-minute exposure time points, while potentially enriched for direct exposure mediators, may fail to capture important sustained or secondary responses, such as oxidative stress or epithelial-to-mesenchymal transformation, which have been suggested as mechanisms that connect exposures with asthma previously (68,73). Finally, our understanding of *SPRY2* was limited to the effects of *SPRY2* knockdown, including its effects on scratch repair. While *SPRY2*-knockdown recapitulated the effect of the rs3861144 risk variant, a better understanding of *SPRY2* biology would require additional experiments, such as *SPRY2* overexpression and a detailed analysis of additional factors that interact with *SPRY2* and ERK1/2 signaling. Our study proposes several avenues for further investigation into this complex but relevant pathway in airway cell physiology and asthma.

Despite these limitations, our study expands the understanding of the genetic factors that influence asthma risk in the context of wildfire PM2.5 exposure, a problem of growing importance due to global climate change. We identified common SNPs that may contribute to the personal risk of asthma associated with wildfire PM2.5 and cellular mechanisms that mediate this risk. We characterized one important mechanism by demonstrating that *SPRY2* expression in response to WSP exposure is mediated by rs3861144, which modifies asthma risk and lung function. This connection between WSP and *SPRY2* may alter asthma risk by impacting airway epithelial cell proliferation and injury repair through the MAPK pathway or increasing pro-inflammatory responses to exposures. Irrespective of the precise mechanism, our results implicate *SPRY2* as a mediator of the adverse effects of wildfire PM2.5 exposure on asthma risk and define *SPRY2* as a potential therapeutic target for asthma.

## Data Availability

All data produced in the study are available on request to the submitting author. All genomic data have been posted for access at the Gene Expression Omnibus with accession number provided in the manuscript.

