## Supplement for "Functional Variant Discovery identifies a novel genetic link between SPRY2, wood smoke, and asthma"

Title

Additional Methods – E1 – E14

E1. Precision Run-on Sequencing (PRO-seq) and Micrococcal Nuclease Chromosome Conformation Capture (Micro-C) data sets

We previously published the Beas-2B PRO-seq nascent transcript dataset (1) used in the current study with linked Gene Expression Omnibus (GEO) accession number GSE167371. Computational analysis to identify transcriptional regulatory elements (TREs) that respond to WSP was performed as described (1) using Tfit (v. 1.0) (2) to annotate the characteristic bidirectional signatures of TREs and define centers of TRE activity. We accessed previously published Micrococcal Nuclease Chromosome Conformation Capture (Micro-C) data from unstimulated Beas-2B cells from GSE241294 for analysis in this study (3).

E2. Public GWAS Data Analysis

We used the Open Targets Genetics online platform (4,5) and the Lung Disease Knowledge Portal (6) to navigate aggregated genetic data, drawing primarily from two GWAS studies of pulmonary physiology (7,8). We performed 52 independent queries in each database and used a padj < 0.05 (Bonferroni) threshold for significance of association. We also performed 52 queries to identify genes associated with genetic variants using the Open Targets Genetics platform expression quantitative trait locus data collection.

E3. Statistical Fine Mapping

Statistical fine mapping at each of the 52 WSP-SNP loci was performed using the Bayesian FINEMAP approach (9), summary statistics from a previous GWAS of forced vital capacity (FVC) (7), linkage disequilibrium maps from the 1000 Genome Project Phase 3 (10), and the R package echolocatoR (11).

E4. Putative Gene-SNP association using shared nascent transcription kinetics in PRO-seq data set

We developed a custom pipeline to assign putative gene targets to each WSP-SNP by applying the Activity-By-Contact paradigm (12) to nascent transcriptional kinetics from PRO-seq. WSP-SNPs were placed in one of 6 categories based on the transcriptional kinetics of their resident TRE – increased or decreased transcription between pairs of experimental conditions (vehicle/control, WSP exposure 30 minutes, WSP exposure 120 minutes). One SNP was allowed to occupy multiple categories. We then collected differentially transcribed genes residing within a 500 kb genomic radius of each SNP in each category and combined these 6 sets (replicates removed) for total aggregate analysis.

E5. Cell Culture, Primary Cells and Reagents

Beas-2B and primary small airway epithelial cell culture were performed as described previously (1,13). Primary small airway cell genotype was determined using a quantitative PCR assay (TaqMan). WSP were obtained from and analyzed by Dr. Andrew Ghio (14,15) and used in cell culture by our group as described previously (1). Live airway epithelial cells were obtained from individuals undergoing research bronchoscopy at the Rocky Mountain Regional Veterans Affairs Medical Center. Informed consent included provisions for using deidentified samples in general research. Cells were collected using cytology brushes, stored in cold saline (0.9% NaCl), and processed within 2 hours of harvest. Primary Human Airway Smooth Muscle Cells were cultured and exposed to TNF-alpha (30 ng/ml final concentration) as described previously (16).

E6. Transcription Factor Binding Motif Predictions and Chromatin Accessibility and Genomic Feature Overlap

Transcription factor binding motif search was performed using the FIMO operation from the MEME suite (v5.0.5) and the HOCOMOCOv11 transcription factor binding motif library (17) in (+/-) 500 bp regions surrounding each WSP-SNP with a statistical threshold of padj < 1e-5 (Benjamini-Hochberg). Genomic overlap between WSP-SNPs and regions of chromatin accessibility defined by ATAC-seq in live airway cells or genomic features (hg38 annotations downloaded from the University of California Santa Cruz genome browser on September 26, 2019) was performed using the intersectBed operation from Bedtools (v2.29.2).

E7. The Assay for Transposase Accessible Chromatin with Sequencing (ATAC-seq)

ATAC-seq was performed as described previously (1,13) using an adaptation of the Omni-ATAC-seq protocol (18). 50e3 cells mechanically dissociated from cytology brushes were processed in duplicate or quadruplicate from each subject. Uniquely indexed libraries (Nextera) were sequenced on an Illumina NovaSeq (University of Colorado Shared Genomics Resource) using paired end sequencing with 150 bp reads. Sequencing data was processed and visualized as described previously (1,13). Raw fastq reads and non-normalized hg38-mapped reads in bedGraph format have been submitted to GEO (GSE283113).

E8. Luciferase assay in plasmid-derived reporter system

Plasmid construction, transfection, and luciferase assays were performed as described previously (20,25). Briefly, we designed PCR primers to amplify 700-800 bp regions surrounding each SNP using Integrated DNA Technologies (IDT) online tools and performed allele change using the QuikChange II Site-directed Mutagenesis (SDM) kit from Agilent. The pMT2A positive control construct has been described previously (25). Experimental plasmids were co-transfected using Lipofectamine 2000 (Life Technologies) with an internal control Renilla luciferase vector (Promega). Luciferase activity was measured using the Dual-Luciferase Reporter Assay System (Promega) and an Infinite M1000 Plate Reader (Tecan). Each experiment was performed in biological quadruplicate and repeated at least once. Statistical testing and p values were computed using two-tailed t-tests. Primer sequences are provided in Supplemental Table E1.

E9. Gene knockdown using short interfering RNA (siRNA) transfection

siRNA studies were conducted using the ON-TARGETplus SMARTpool against *SPRY2* (*SPRY2*-siRNA, L-005206-00-0005) or non-targeting control (siCTRL, D-001810-10-20) from Dharmacon. Beas-2B cells were transfected with 50 nM siRNA (Dharmacon) using the Lipofectamine RNAiMAX transfection reagent according to the manufacturer's protocol (Life Technologies) as described previously (19). Twenty-four to 48 hours later, cells were lysed for RNA or protein analysis or subjected to scratch assay.

E10. RNA extraction, RNA-sequencing, and Quantitative PCR

Quantitative PCR (qPCR) and RNA extraction were performed as described previously (1,20). Briefly, bulk RNA was extracted from cells lysed in Trizol (Life Technologies) using the RNA PureLink kit. Following quality control analysis, RNA was submitted for bulk sequencing.

RNA sequencing libraries were prepared according to the Watchmaker mRNA Library Build user guide. Briefly, mRNA from 200 ng of total RNA was isolated using polyA, oligo-dT magnetic beads. The isolated mRNA was then subject to enzymatic fragmentation, resulting in ~200 bp fragments. The resulting RNA fragments underwent first and second strand cDNA synthesis. IDT xGen Stubby Adapters were then ligated to the cDNA. The ligated product was then PCR amplified for 10 cycles using the IDT xGen UDI Primers. The resulting libraries were quantified using the Qubit hsDNA assay and TapeStation DNA 1000 assay. Equal molar concentrations were pooled, diluted to 800 pM, and sequenced using a 200 cycle Illumina NextSeq 2000 flow cell.

We also performed reverse transcription and qPCR on purified RNA. All qPCR results are presented as normalized to the *RPL19* gene. Statistical analysis and computation of p values for qPCR experiments were performed using two-tailed t-tests of unpaired samples with equal distributions (LibreOffice v24.8.6.2). SYBR Green primer sequences for qPCR experiments are provided in Supplemental Table E3.

E11. Bioinformatic analysis of RNA-sequencing data

RNA sequencing data in fastq format was processed and mapped to the genome as described previously (1,13) with minor adjustments for RNA data, such as allowing for spliced alignments in the hisat2 operation. Alignment rates were > 95% for all libraries. Gene reads were then counted using featureCounts in the Rsubread package (21), and differential expression analysis was performed using DESeq2 (22) for pairwise comparisons. Raw fastq reads, non-normalized reads in bedGraph format, and raw count data from featureCounts (non-normalized) have been submitted to GEO (GSE283113).

E12. Western Blot

Western blotting was performed to detect protein concentrations according to established protocols (19). Given overlapping molecular sizes of proteins of interest, stripping buffer (1% Sodium dodecyl sulfate and 2M glycine, pH 2.2) was applied to probe multiple targets from the same blot. All Western Blot experiments were repeated twice with qualitatively similar results, and the best image was selected for presentation. In an attempt to increase authenticity, complete blot images (with additional information not pertinent to the primary analysis) are provided in Supplemental Figures E3-E6. Antibodies used for Western Blot included anti-*SPRY2* S1444 (Sigma), anti-beta-tubulin ab52901 (Abcam), anti-phospho-ERK1/2 4370 (Cell Signaling), anti-ERK1/2 4695 (Cell Signaling), anti-GAPDH sc-25778 (Santa Cruz), anti-Lamin-B1 12586S (Cell Signaling) horseradish-peroxidase-conjugated anti-rabbit NA93401ML (GE Healthcare/Amersham), horseradish-peroxidase-conjugated anti-mouse NA931 (GE Healthcare/Amersham).

E13. Scratch Assay

Following transfection with siRNA, Beas-2B cells were grown to a confluent monolayer, a mechanical scratch was made using a P1000 pipette tip, and photographs were taken at multiple time points. Quantification of scratch closure was performed using ImageJ software (23) and normalized as a fraction of the scratch area at the 0 hour time point. Scratch outlines were constructed using an online tool (24) and then manually reviewed for each image to ensure accurate scratch border representation. The scratch area was computed by ImageJ following confirmation of the scratch border in pixel-based units and then normalized to initial scratch area of each well. Any scratch with complete closure within 12 hours was excluded, as this resulted in wells with small and incomplete scratches. Statistical testing was performed using a two-tailed t-test to compare means between control and *SPRY2* knockdown groups. Multiple testing for multiple time points was accounted for using the Bonferroni correction method. This experiment was performed in biologic quadruplicate and repeated 4 times.

E14. Interleukin-8 Quantification by Enzyme linked Immunosorbent Assay (ELISA)

Quantification of IL-8 was performed by a commercial ELISA kit (R&D Systems). Briefly, plates were prepared with anti-IL-8 capture antibody prior to the addition of sample. Cell supernatant was harvested following exposure and applied directly to prepared plates. Anti-IL-8 detection antibody was then added, and IL-8 levels were quantified from streptavidin-Horseradish peroxidase associated with the detection antibody using absorbance read by an Infinite M1000 plate reader (Tecan).

Table E1 – Cloning PCR primers used to construct plasmid reporters

| Cloning primer | Sequence |
| --- | --- |
| rs75459144-fw | GGTGGCATCTCTTCACTGTT |
| rs75459144-rv | GGCTCGCAAGTCATAACAAATC |
| rs9903086-fw | GGGATGGAATGTCAGAAGGAAA |
| rs9903086-rv | GGTGGAAAGAGACCTACATTGG |
| rs28362965-fw | GAGGCTGGTGTCTCCTTTATC |
| rs28362965-rv | CTGAAGACCCATAGCATCCAG |
| rs12630368-fw | GATCTTTATAGCTGGGCTGGTC |
| rs12630368-rv | CCTCATCAGTGAAGTGAGTGAAA |
| rs1371799-fw | AGCTGGTCACATTGGCTATATT |
| rs1371799-rv | CCAGGTGATGTTACAGCTCTC |
| rs3739819-fw | CCACTGCTGCAGGTCTTAAA |
| rs3739819-rv | TGTGCTTCTTTGGGTCACTTAT |
| rs6871945-fw | CGAGGGAGGAGCACTAAATA |
| rs6871945-rv | TGAGCCATTCATTCCCAAAG |
| rs11832287-fw | CAGGTCCCAGTCGAGAAA |
| rs11832287-rv | CTCCTACTGTTCTGGTCACA |
| rs9509353-fw | CAACTCCAGCATCGACAAA |
| rs9509353-rv | ATCCAAGGATTCCAATTATGCT |
| rs10424200-fw | GTGTGACTTTCAGGGTTCAG |
| rs10424200-rv | CGACACAGCCAAATGAAATAAG |
| rs10418662-fw | GACTCAGCGTGTAGAGAAAG |
| rs10418662-rv | CCTGAATATGTGTATGGGTATCA |
| rs3861144-fw | GTTTGGTTCGTTGCTCATATAC |
| rs3861144-rv | CAGGCCAGCTTGGAATAG |
| rs11883931-fw | TCACACATTCAACCCACTTT |
| rs11883931-rv | GCCTGAGTCTGCTTAACATC |
| rs1873333-fw | GTGGTCTGGGTTGTGAATC |
| rs1873333-rv | GAATCACACTACCTGCTCTC |
| rs644821-fw | CCACTATAATGCCCAGCTAAA |
| rs644821-rv | CCACAGGGAACATGTGTAAG |
| rs4885681-fw | CAGGCATCTTACAGAATGGG |
| rs4885681-rv | CCATCTGTCCCTGATATCCATTT |
| rs9913846-fw | CGCTTATAATCCCAGCATGT |
| rs9913846-rv | AAGCTCATGGACCACTCT |
| rs9911655-fw | GGATGATCAGCTATGGGTAAAG |
| rs9911655-rv | CTTGTCAGCACTGGGTATTAT |
| rs33914436-fw | GTGATGGGTCCATATTCATAGAG |
| rs33914436-rv | AACTGAGTCTTGTCCGTTTC |
| Site-directed Mutagenesis Primer | Sequence |
| rs3861144_C>A-fw | GTGGAGGATGTGGCTCAGGAAGAGGGAAG |
| rs3861144_C>A-rv | CTTCCCTCTTCCTGAGCCACATCCTCCAC |
| rs11883931_T>G-fw | CAATCGGCTCAAATCCCTGTCCTTATTTGAATTTCAATATAAAAATCCATT |
| rs11883931_T>G-rv | AATGGATTTTTATATTGAAATTCAAATAAGGACAGGGATTTGAGCCGATTG |
| rs9903086_C>T-fw | GTGTTGCTTAGGAAGTTGGTGAACCAGCTCTGGAG |
| rs9903086_C>T-rv | CTCCAGAGCTGGTTCACCAACTTCCTAAGCAACAC |
| rs12630368_G>C-fw | TTTCCGAATGGGTATCCCTGTGCTCTGGAGGG |
| rs12630368_G>C-rv | CCCTCCAGAGCACAGGGATACCCATTCGGAAA |
| rs11832287_A>G-fw | CTGCCACACTCTCCGAAGGGTCTCCCC |
| rs11832287_A>G-rv | GGGGAGACCCTTCGGAGAGTGTGGCAG |

Table E2 – crRNA sequences used to construct Cas9 ribonucleic protein complexes for genome editing and gene knockdown

| crRNA | Sequence |
| --- | --- |
| rs3861144-TRE-5’-crRNA | GCATCCAAAGAGCTAGGCCCAGG |
| rs3861144-TRE-3’-crRNA | GTCAAGGTGCTGAAGACATCTGG |
| SPRY2-5’-crRNA | GTAAGGAGTGCACCTACCCAAGG |
| SPRY2-3’-crRNA | CCTACTGTCGTCCCAAGACCTGG |

Table E3 – PCR and quantitative PCR primers used to confirm genome editing and quantify gene expression

| PCR primer | Sequence |
| --- | --- |
| rs3861144-TRE 5’ PCR | TCCCTGTCCATTCACTTAAAC |
| rs3861144-TRE 3’ PCR | TGAACCTTATGCAAATCCTACT |
| qPCR primer |  |
| SPRY2-fw | TGTGGCAAGTGCAAATGTAAG |
| SPRY2-rv | CAGCATACACAAGTCCCATAGT |
| CYP1A1-fw | GTAGTGCTCCTTGACCATCTTC |
| CYP1A1-rv | GGTGCTATCGACAAGGTGTTA |
| CXCL8-fw | CTTGGCAGCCTTCCTGATTT |
| CXCL8-rv | GGGTGGAAAGGTTTGGAGTATG |

Table E4 – Genes with expression Quantitative Trait Locus associations with WSP-SNPs in the Open Targets Genetics platform

| *ACTN4* | *CCDC92B* | *ENG* | *JUP* | *MICB* | *PIGL* | *RRP12* | *TRIM26* |
| --- | --- | --- | --- | --- | --- | --- | --- |
| *ADORA2B* | *CCHCR1* | *FKRP* | *KIF25* | *MINK1* | *PIP5KL1* | *SERP2* | *TRPV2* |
| *AFDN* | *CD9* | *FLOT1* | *KLF11* | *MIR3681HG* | *PLCL1* | *SF3B1* | *TTC16* |
| *AK1* | *CDK9* | *FPGS* | *KRT23* | *MPPE1* | *PLD2* | *SFRP5* | *TTC19* |
| *AMH* | *CENPV* | *FSTL3* | *KRT32* | *NCOR1* | *PLEKHD1* | *SH2D3C* | *TTI1* |
| *AMOTL1* | *CFAP157* | *G0S2* | *KRT34* | *NDUFB3* | *PLEKHJ1* | *SIRT2* | *UBB* |
| *ANKRD44* | *CFLAR* | *GALNT6* | *LACC1* | *NECAB2* | *PNMA8A* | *SLC25A25* | *VARS2* |
| *ANKRD55* | *CHRNE* | *HAP1* | *LAMB3* | *NIBAN2* | *PNMA8B* | *SLC4A8* | *VSTM2L* |
| *ANXA10* | *CLDN1* | *HK2* | *LAMC1* | *NIF3L1* | *PNMA8C* | *SORBS2* | *VWF* |
| *ARHGAP19* | *CLK1* | *HLA-C* | *LAMC2* | *NMNAT2* | *PPIL3* | *SPRY2* | *XPO4* |
| *ARPC5* | *CNP* | *HLA-E* | *LARGE1* | *NRM* | *PPP5C* | *ST6GALNAC4* | *ZFYVE27* |
| *AVPI1* | *COL6A3* | *HLA-G* | *LGALS4* | *NT5C3B* | *PRKCA* | *ST6GALNAC6* | *ZNF232* |
| *BHLHE40* | *COMTD1* | *HTR1B* | *LGALS7* | *ODAD4* | *PRKD2* | *STIM2* | *ZNF385C* |
| *BOLL* | *CPNE9* | *HYCC2* | *LGALS7B* | *ORC2* | *PRR16* | *STXBP1* | *ZSWIM7* |
| *C17orf107* | *CXCL2* | *IER3* | *LOC105377862* | *OSGIN1* | *PRSS57* | *SUSD6* |  |
| *CAMTA2* | *CXCL5* | *IFT88* | *LPAR1* | *P3H2* | *PTRH1* | *TBC1D19* |  |
| *CAPN12* | *CYS1* | *IL17D* | *LRMDA* | *P3H4* | *RFTN2* | *TBL1XR1* |  |
| *CASP10* | *DDX60L* | *IL1RAP* | *LRRFIP1* | *PDLIM3* | *RINL* | *TGM2* |  |
| *CASP8* | *EEF1AKMT1* | *IMPA2* | *MARS2* | *PEAK3* | *RPRD1B* | *TP63* |  |
| *CCDC8* | *EEIG1* | *JSRP1* | *MARVELD1* | *PI4K2A* | *RRN3P2* | *TRIB2* |  |

Table E5 – Putative gene targets of WSP-SNPs identified from shared nascent transcript kinetics and location within 500 kb

| *ABCA7* | *CASP10* | *ENO3* | *IL31RA* | *LOC101929882* | *ODF2* | *SGSM2* | *TPGS1* |
| --- | --- | --- | --- | --- | --- | --- | --- |
| *ABCF1* | *CASP8* | *EPGN* | *IL6ST* | *LOC102723566* | *OGG1* | *SH2B1* | *TRAK2* |
| *ACLY* | *CCDC122* | *EREG* | *INCA1* | *LOC105371998* | *ORC2* | *SH2D3C* | *TRIB2* |
| *ACP7* | *CCDC144A* | *EXD2* | *ITPR1* | *LOC105377590* | *OSGIN1* | *SHC2* | *TRIM39* |
| *ACRBP* | *CCDC61* | *FAM102A* | *JAGN1* | *LOC339975* | *P3H2* | *SIRT2* | *TRIM39-RPP21* |
| *AFDN* | *CCDC8* | *FAM126B* | *JUP* | *LOC388242* | *P3H4* | *SLC25A11* | *TSC22D1* |
| *AK1* | *CCDC92B* | *FAM149A* | *KAT2A* | *LOC613038* | *PALM* | *SLC25A25* | *TTC16* |
| *ALOX15* | *CD9* | *FAM98C* | *KAT6B* | *LPAR1* | *PANX1* | *SLC25A25-AS1* | *TTI1* |
| *AMH* | *CDC34* | *FBXO17* | *KCNK6* | *LRMDA* | *PDLIM3* | *SLC27A4* | *TTLL3* |
| *AMOTL1* | *CDK9* | *FBXO27* | *KIF1C* | *LRRFIP1* | *PIP5KL1* | *SLC35G4* | *TUBB* |
| *ANKRD37* | *CEP112* | *FGF22* | *KLHL11* | *LRSAM1* | *PLCL1* | *SLC39A3* | *TUFM* |
| *ANKRD55* | *CERCAM* | *FKBP10* | *KRT15* | *LTBR* | *PLD2* | *SLX1A* | *UBE2G1* |
| *ANO2* | *CFAP157* | *FLACC1* | *KRT16* | *MADCAM1* | *PLEKHJ1* | *SLX1A-SULT1A3* | *UBTD1* |
| *ANXA10* | *CFD* | *FLJ31104* | *KRT17* | *MARVELD1* | *PLPP2* | *SLX1B* | *UFSP2* |
| *AOX1* | *CHMP1B* | *FLJ38576* | *KRT19* | *MDC1* | *POLR2E* | *SLX1B-SULT1A4* | *URM1* |
| *AREG* | *CIZ1* | *FLOT1* | *KRT32* | *MED16* | *POLRMT* | *SMG1P2* | *UTP25* |
| *ARHGAP19* | *CLDN1* | *FPGS* | *KRT33B* | *MIER2* | *PPIL3* | *SMIM2* | *VAMP1* |
| *ARHGAP19-SLIT1* | *CLK1* | *FRAT1* | *KRT34* | *MINK1* | *PPP1R10* | *SMIM2-AS1* | *VMO1* |
| *ARHGAP40* | *CLUH* | *FRAT2* | *KRTAP2-1* | *MIR1268A* | *PPP1R18* | *SMIM2-IT1* | *VWF* |
| *ARHGAP45* | *CNN2* | *FSTL3* | *KRTAP2-2* | *MIR219B* | *PPP5C* | *SNORA65* | *WDR18* |
| *ARID3A* | *CNP* | *FUT4* | *KRTAP2-3* | *MIR2861* | *PRELID3A* | *SNORD49B* | *ZMYND15* |
| *ARL8B* | *COL6A3* | *GALNT16* | *KRTAP2-4* | *MIR3187* | *PRKCA* | *SNORD65* | *ZNF232* |
| *ARPC4* | *COMTD1* | *GAPDH* | *KRTAP4-6* | *MIR3680-1* | *PRKD2* | *SNX25* | *ZNF287* |
| *ARPC4-TTLL3* | *COPS8* | *GARNL3* | *KRTAP4-7* | *MIR3680-2* | *PRR16* | *SPAG7* | *ZNF384* |
| *ARPC5* | *CPNE9* | *GHDC* | *KRTAP4-8* | *MIR3681HG* | *PRR3* | *SPIRE1* | *ZNF503* |
| *ARRB2* | *CRELD1* | *GLTPD2* | *KRTAP4-9* | *MIR3960* | *PRRT3* | *SPNS1* | *ZNF503-AS2* |
| *ATAT1* | *CSNK1G2* | *GNAL* | *KRTAP9-7* | *MIR4745* | *PRRT3-AS1* | *SPPL2B* |  |
| *ATP2A1* | *CTNNBL1* | *GNL1* | *LACC1* | *MIR6789* | *PTBP1* | *SPRED3* |  |
| *ATP2C2-AS1* | *CWC15* | *GOLGA2* | *LAMC1-AS1* | *MIR877* | *PTGES2* | *SPRY2* |  |
| *ATP8B3* | *CXCL1* | *GPX4* | *LAMC2* | *MISP* | *PTGES2-AS1* | *ST6GALNAC4* |  |
| *ATXN2L* | *CXCL16* | *GRHL1* | *LARGE1* | *MLYCD* | *PTRH1* | *ST6GALNAC6* |  |
| *AUP1* | *CXCL2* | *GZMM* | *LAT* | *MNT* | *R3HCC1L* | *STAT5A* |  |
| *AVPI1* | *CXCL3* | *HAP1* | *LHFPL4* | *MORN4* | *R3HDM4* | *STAT5B* |  |
| *BHLHE40* | *CXCL8* | *HCG17* | *LINC00243* | *MPPE1* | *RAB17* | *STRADB* |  |
| *BHLHE40-AS1* | *CYP4V2* | *HCG20* | *LINC00390* | *MRPL51* | *RAB5C* | *STXBP1* |  |
| *BOLA2* | *CYS1* | *HCG27* | *LINC00578* | *MRPL57* | *RABEP1* | *SULT1A3* |  |
| *BOLA2B* | *DAZAP2* | *HCN2* | *LINC01002* | *MTHFD2L* | *RABEP2* | *SULT1A4* |  |
| *BOLA2-SMG1P6* | *DDX60* | *HCRT* | *LINC01170* | *MUSK* | *RALGAPB* | *SUSD6* |  |
| *BRPF1* | *DDX60L* | *HK2* | *LINC01255* | *MYBBP1A* | *RINL* | *SVEP1* |  |
| *BSG* | *DHX16* | *HLA-E* | *LINC01291* | *NCF2* | *RNF126* | *SWI5* |  |
| *BTBD2* | *DHX58* | *HPCAL1* | *LINC01698* | *NDFIP2* | *RPP21* | *TAF1C* |  |
| *BZW1* | *DIRAS1* | *HSBP1* | *LINC01775* | *NDUFB3* | *RPRD1B* | *TAPBPL* |  |
| *C17orf107* | *DNAAF1* | *HSDL1* | *LINC02015* | *NECAB2* | *RPUSD3* | *TBC1D19* |  |
| *C1orf74* | *DNAJC7* | *HTR1B* | *LINC02492* | *NFATC2IP* | *SAMD8* | *TFCP2* |  |
| *C2CD6* | *DNM1* | *IER3* | *LINC02570* | *NIBAN2* | *SBNO2* | *TGM2* |  |
| *C5orf67* | *DQX1* | *IER3-AS1* | *LMNB2* | *NIF3L1* | *SCAMP4* | *TIMM13* |  |
| *C9orf16* | *EDEM1* | *IFFO1* | *LOC100288123* | *NKIRAS2* | *SCARNA10* | *TLR3* |  |
| *CALM3* | *EGOT* | *IL17D* | *LOC100505782* | *NMNAT2* | *SCN8A* | *TMEM259* |  |
| *CAMK1* | *EIF1* | *IL17RC* | *LOC100996750* | *NOP2* | *SERP2* | *TNFRSF1A* |  |
| *CAMTA2* | *ENG* | *IL17RE* | *LOC101929165* | *NT5C3B* | *SGO2* | *TOR2A* |  |

Table E6 – Statistical fine mapping results at 7 WSP-SNP loci with non-zero credible set associated with Forced Vital Capacity

| WSP-SNP | SNPs (Credible Set) | Posterior Probability |
| --- | --- | --- |
| rs6584144 | rs4919120 | 1 |
|  | rs114106004 | 1 |
|  | rs111439614 | 1 |
|  | rs111393981 | 1 |
|  | rs10444067 | 1 |
| rs9509353 | rs9578309 | 1 |
|  | rs9550665 | 1 |
|  | rs3000678 | 1 |
|  | rs2772175 | 1 |
|  | rs2442454 | 1 |
| rs9939094 | rs7192141 | 1 |
|  | rs62038226 | 1 |
|  | rs252253 | 1 |
|  | rs11862473 | 1 |
|  | rs114961054 | 1 |
| rs9903086 | rs9913846 | 1 |
|  | rs9911655 | 1 |
|  | rs33914436 | 1 |
|  | rs11869280 | 1 |
|  | rs113146141 | 1 |
| rs11590328 | rs79813470 | 1 |
|  | rs6689266 | 1 |
|  | rs6689166 | 1 |
|  | rs189117755 | 1 |
|  | rs112931592 | 1 |
| rs6784916 | rs567321108 | 1 |
|  | rs566280492 | 1 |
|  | rs192351294 | 1 |
|  | rs141524859 | 1 |
|  | rs12490194 | 1 |
| rs3129975 | rs77645489 | 1 |
|  | rs4713354 | 1 |
|  | rs3095330 | 1 |
|  | rs3094118 | 1 |
|  | rs116473430 | 1 |

Table E7 – Functional Annotation Categories for Genes upregulated or downregulated in *SPRY2* knockdown-WSP exposed (si*SPRY2*-WSP) cells compared with control-WSP exposed (siCTRL-WSP) cells.

| si*SPRY2*-WSP2h > siCTRL-WSP2h |  |  |
| --- | --- | --- |
| **Term** | **Enrichment score** | **FDR** |
| Endoplasmic Reticulum | 2.7 | 0.012 |
| Synapse | 1.9 | 0.022 |
| TGF-beta | 1.9 | 0.017 |
| Glycoprotein | 1.8 | 0.022 |
| si*SPRY2*-WSP2h < siCTRL-WSP2h | |  |
| **Term** | **Enrichment score** | **FDR** |
| Metalloprotease | 2.1 | 0.037 |
| Kinase | 1.9 | 0.0062 |
| Transmembrane | 1.8 | 0.003 |
| si*SPRY2*-WSP4h > siCTRL-WSP4h |  |  |
| **Term** | **Enrichment score** | **FDR** |
| Growth Factor | 2.5 | 0.028 |
| Secreted | 2.1 | 0.027 |
| TNF signaling pathway | 1.8 | 0.023 |
| si*SPRY2*-WSP4h < siCTRL-WSP4h | |  |
| **Term** | **Enrichment score** | **FDR** |
| None |  |  |

Table E8 – Genes upregulated in Beas-2B control cells (siCTRL) at 4 hours of WSP exposure compared with 2 hours of exposure. Genes in bold are not identified in the corresponding *SPRY2* knockdown gene set (Table E8). Genes without an official gene symbol (n = 17) have been removed from this list.

| siCTRL WSP4h > WSP2h | | |  |  |  |
| --- | --- | --- | --- | --- | --- |
| *ADAMTS1* | *DIO2* | *IL4R* | *MYPN* | *PTHLH* | ***TMEM156*** |
| *ANGPTL4* | *DKK1* | *INAVA* | *NFATC1* | *PTPRE* | ***TMEM171*** |
| *APCDD1L* | *DPF3* | *INHBA* | *NFATC2* | *PTX3* | *TNFRSF9* |
| *ARHGAP31* | *DSEL* | *INSYN2B* | *NHS* | *RIMBP3* | *TNFSF15* |
| *ARHGEF16* | *DUSP10* | ***ISL1*** | *NLRP3* | ***RIMBP3B*** | *TRAF1* |
| *ATP9A* | *ESM1* | *ITGA2* | *NMRAL2P* | ***RIMBP3C*** | *VEGFC* |
| *BACH2* | *FAM167A-AS1* | ***IVL*** | *NPAS1* | ***RNF165*** | *VEPH1* |
| *CARD6* | *FBXO32* | *KCNG1* | ***P2RY6*** | *ROR1* | *WTAPP1* |
| ***CCIN*** | ***FOLR3*** | *KCNJ12* | ***PCBP3*** | *S1PR3* | *ZHX2* |
| *CHAC1* | *GRIN2A* | ***KLF17*** | *PCDH9* | ***SCAT1*** | *ZNF365* |
| *CHAD* | *HDAC9* | *MAML2* | *PDE4B* | *SERPINB10* | *ZNF737* |
| ***CHRM3*** | ***HDHD3*** | *MMP1* | *PGM2L1* | *SERPINB2* |  |
| ***CHST2*** | *HECW2* | *MMP3* | *PMEPA1* | *SESN2* |  |
| *COBL* | *HMOX1* | *MMP9* | ***PROX2*** | *SHISAL1* |  |
| *CSF2* | *IL32* | *MSC* | *PTGFR* | *TANC2* |  |

Table E9 – Genes upregulated in *SPRY2* knockdown Beas-2B cells at 4 hours of WSP exposure compared with 2 hours of exposure. Genes in bold are not identified in the corresponding control gene set (Table E7). Genes without an official gene symbol (n = 23) have been removed from this gene list.

| si*SPRY2* WSP4h > WSP2h | | |  |  |  |
| --- | --- | --- | --- | --- | --- |
| ***ACTC1*** | *CHAD* | *HECW2* | ***LAMC2*** | *PMEPA1* | ***SSTR2*** |
| *ADAMTS1* | ***CITED4*** | ***HEYL*** | ***LINC00942*** | ***PRKG1-AS1*** | ***ST3GAL1*** |
| ***ADGRG1*** | ***CLDN1*** | ***HHIP*** | ***LINC02454*** | *PTGFR* | ***TAF4B*** |
| ***AMPD3*** | *COBL* | ***HLX*** | ***LRRC8C*** | *PTHLH* | *TANC2* |
| *ANGPTL4* | *CSF2* | *HMOX1* | *MAML2* | *PTPRE* | *TEX26-AS1* |
| ***ANXA10*** | ***CXCL3*** | ***HSD11B1-AS1*** | ***MAP3K4*** | *PTX3* | ***TFPI2*** |
| *APCDD1L* | ***CXCL5*** | ***ICAM1*** | ***MEDAG*** | ***RASSF10*** | ***TLR2*** |
| ***APCDD1L-DT*** | ***CXCL8*** | ***IGFN1*** | *MMP1* | ***RELB*** | ***TNFAIP3*** |
| *ARHGAP31* | *DIO2* | ***IL1A*** | *MMP3* | *RIMBP3* | *TNFRSF9* |
| *ARHGEF16* | *DKK1* | ***IL1B*** | *MMP9* | ***ROBO4*** | *TNFSF15* |
| *ATP9A* | ***DOCK2*** | ***IL27RA*** | *MSC* | *ROR1* | ***TNFSF8*** |
| *BACH2* | *DPF3* | *IL32* | ***MSC-AS1*** | ***RRAD*** | *TRAF1* |
| ***BBS12*** | ***DRAM1*** | *IL4R* | *MYPN* | *S1PR3* | ***TSPAN18*** |
| ***BCL2A1*** | *DSEL* | *INAVA* | ***NAV3*** | ***SAMD9*** | ***VCAM1*** |
| ***BEGAIN*** | *DUSP10* | *INHBA* | *NFATC1* | *SERPINB10* | *VEGFC* |
| ***BIRC2*** | ***ENTPD7*** | ***INHBA-AS1*** | *NFATC2* | *SERPINB2* | *VEPH1* |
| ***BIRC3*** | *ESM1* | *INSYN2B* | ***NFKB2*** | *SESN2* | *WTAPP1* |
| ***BTG1*** | ***EVA1C*** | ***IRAK2*** | *NHS* | ***SGK1*** | *ZHX2* |
| ***C1QTNF1*** | ***FAM167A*** | ***ITGA11*** | *NLRP3* | *SHISAL1* | ***ZMIZ1*** |
| ***C1QTNF1-AS1*** | *FAM167A-AS1* | *ITGA2* | *NMRAL2P* | ***SLC12A5-AS1*** | *ZNF365* |
| ***C6orf141*** | ***FAP*** | ***KCNF1*** | *NPAS1* | ***SLC41A2*** | ***ZNF491*** |
| *CARD6* | *FBXO32* | *KCNG1* | ***NRG1*** | ***SMAD9*** | *ZNF737* |
| ***CARD8-AS1*** | ***G0S2*** | *KCNJ12* | ***OSGIN1*** | ***SMOX*** |  |
| ***CARMIL2*** | ***GEM*** | ***KCNJ18*** | ***PAQR9*** | ***SOD2*** |  |
| ***CCNE1*** | ***GNGT1*** | ***KRT23*** | *PCDH9* | ***SOD2.1*** |  |
| ***CDC14A*** | *GRIN2A* | ***L3MBTL3*** | *PDE4B* | ***SP6*** |  |
| *CHAC1* | *HDAC9* | ***LAMB3*** | *PGM2L1* | ***SPHK1*** |  |

Figure E1 – Statistically fine mapped variants at the rs9903086/*JUP* locus

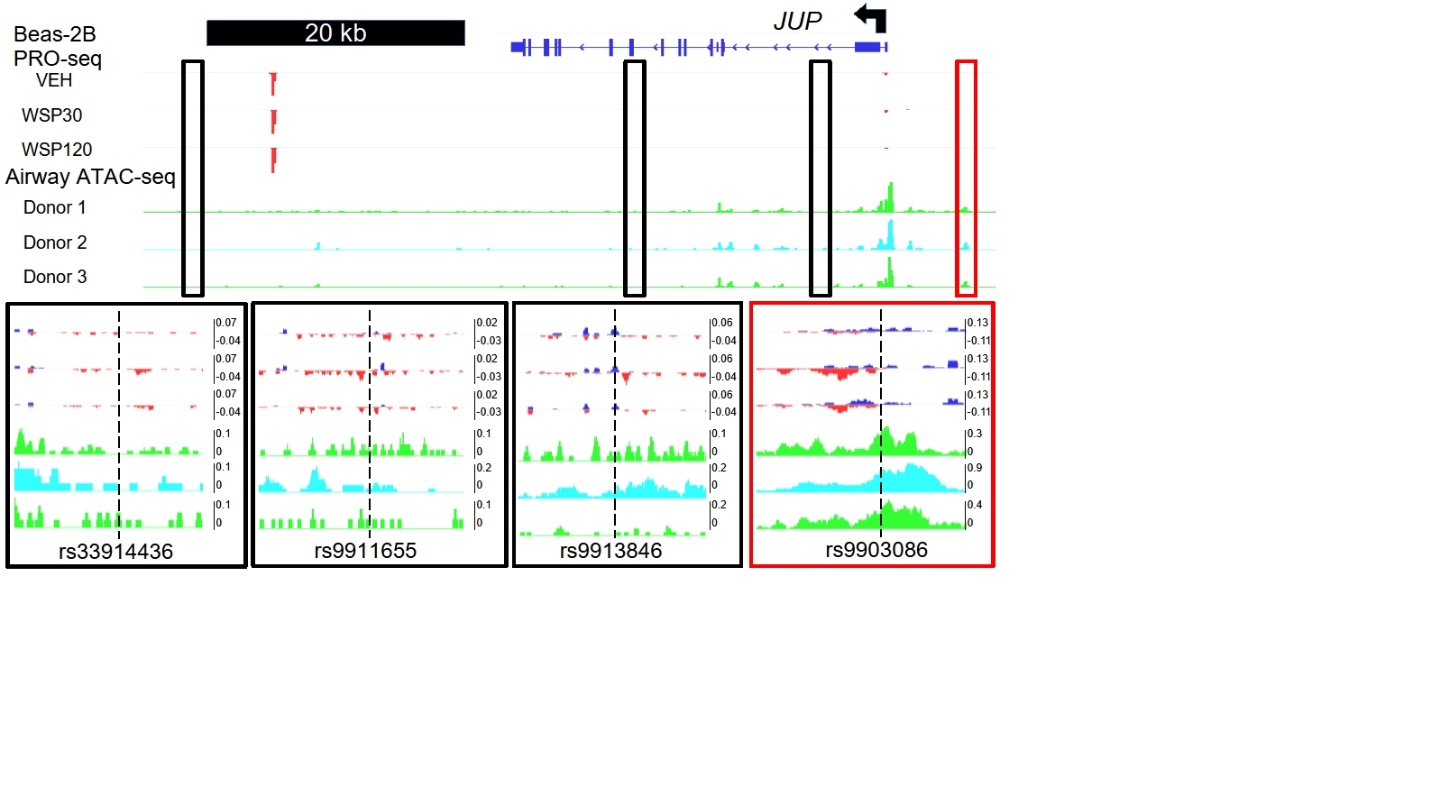

Figure E2 – Micrococcal Nuclease Chromosome Conformation Capture (Micro-C) shows that rs3861144 and *SPRY2* reside within a shared topologically associated domain and interact indirectly

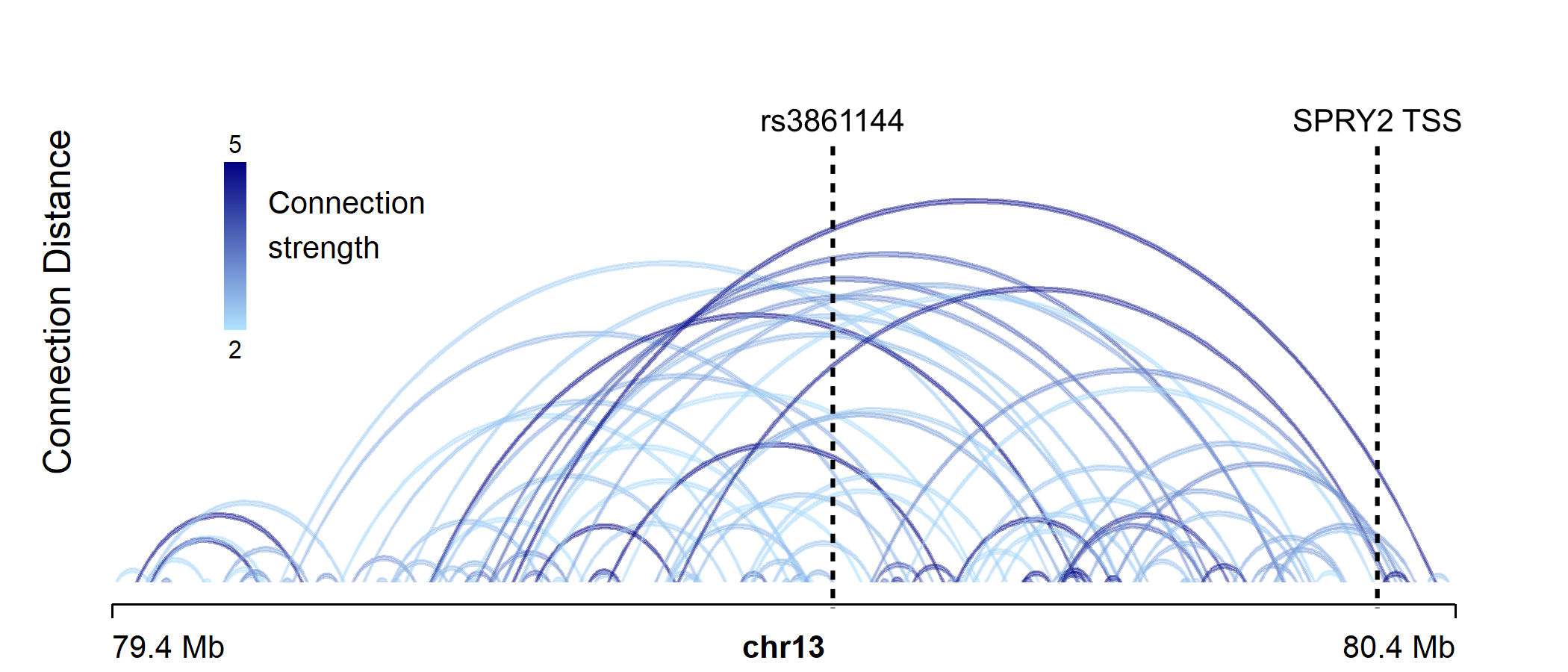

Figure E3 – Beas-2B cells exposed to WSP exhibit an increase in *SPRY2* expression in a dose dependent fashion. Left - qPCR demonstrates dose dependence. Right - Western blot demonstrates an increase in *SPRY2* expression in Beas-2B cells following WSP exposure for 24 hours (WSP 24h); GAPDH served as the loading control.

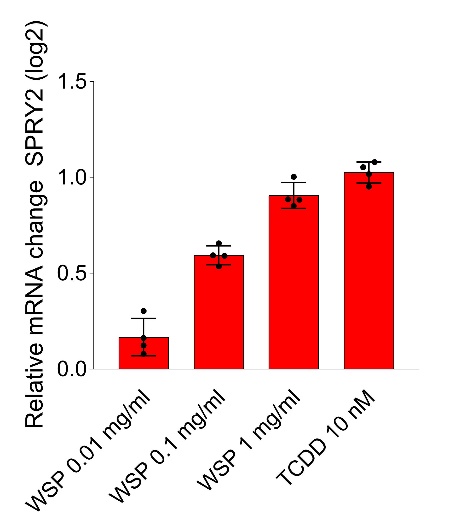

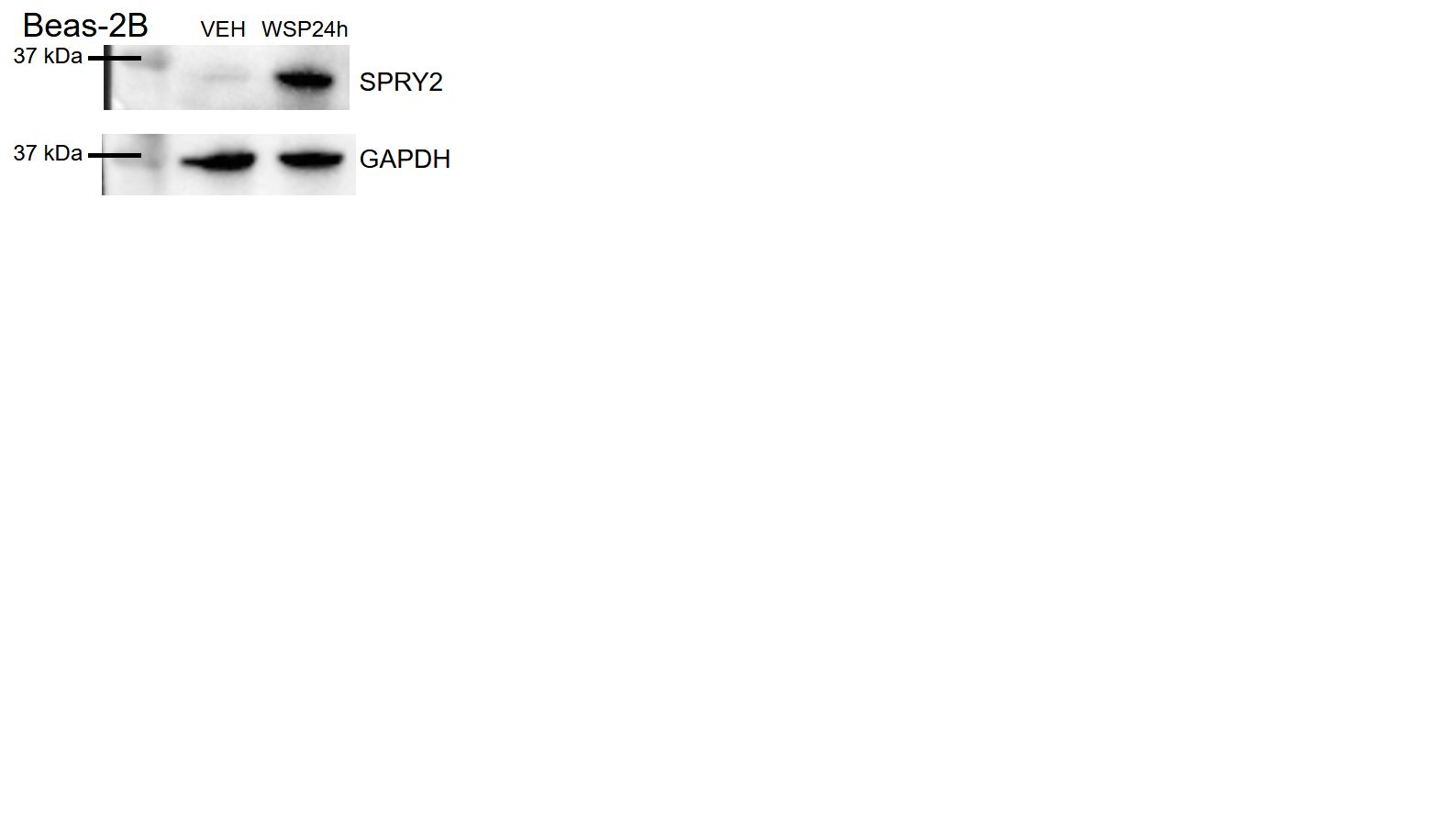

Figure E4 – Principal Component Analysis of RNA sequencing data. VEH = vehicle control cells; WSP = Wood Smoke Particle exposure for 2 or 4 hours; siCTRL = scrambled control transfected cells; si*SPRY2* = cells transfected with *SPRY2*-targeting siRNA.

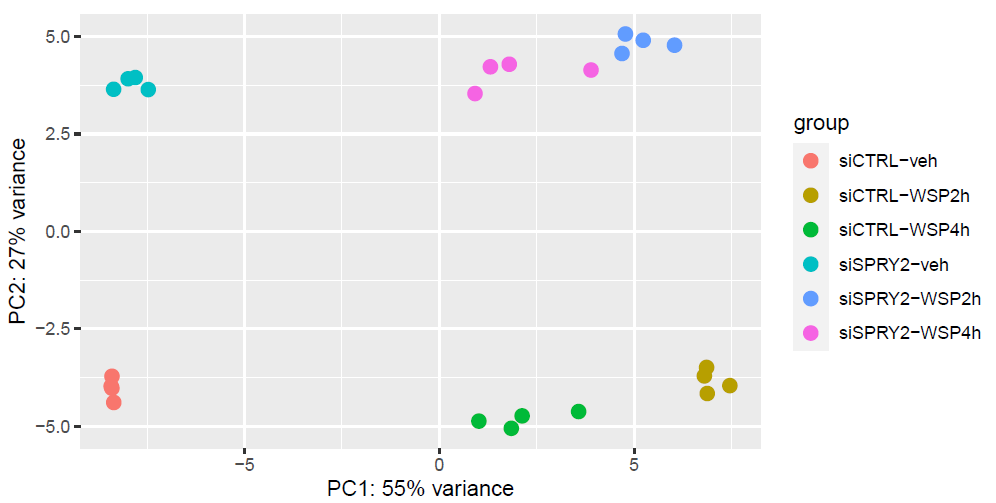

Figure E5 –Western Blot demonstrates a change in phosphorylated ERK1/2 kinetics following acute WSP exposure in Beas-2B cells with *SPRY2* knockdown (si*SPRY2*). A portion of this image was used in Figure 6C.

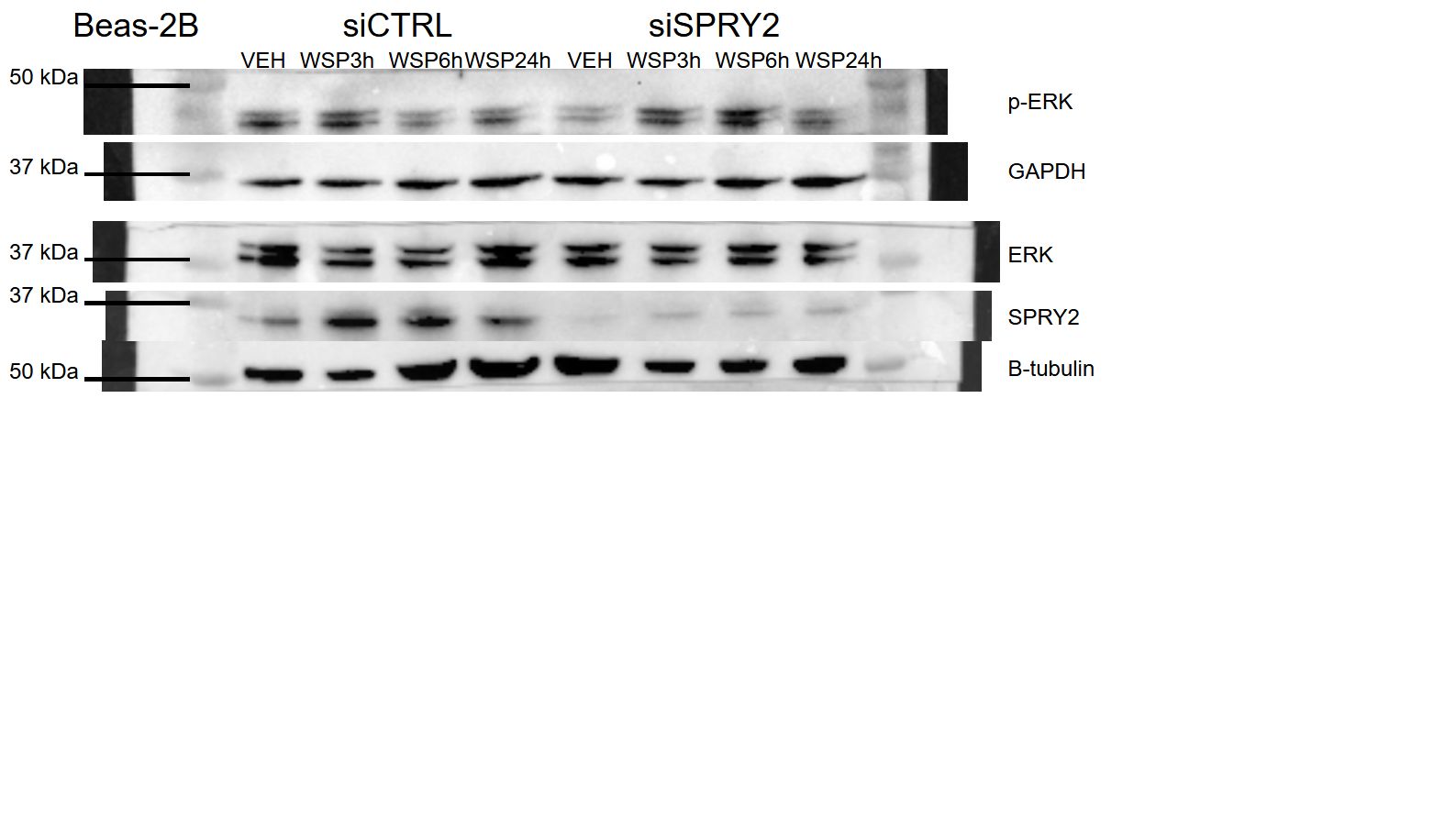

Figure E6 – CXCL8 gene expression is reduced in si*SPRY2* cells following 24 hours of WSP exposure.

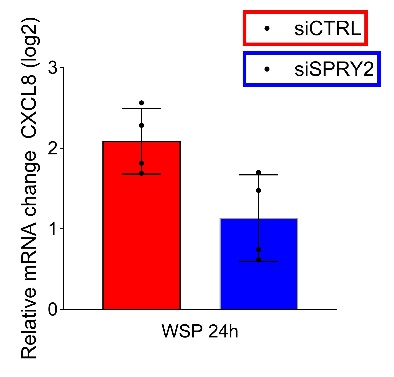

Figure E7 – LDH release assay in cell media from control and *SPRY2*-knockout Beas-2B cells showed no significant change in LDH release or cytotoxicity with WSP exposure at 24 hours.

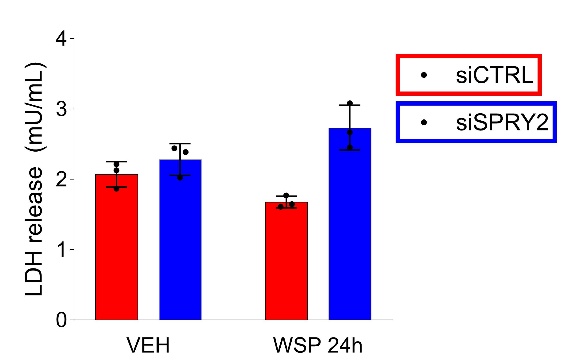

Figure E8 – Full Western Blot images of HASM cells from normal/healthy and asthmatic donors exposed to TNF-alpha shown in main Figure 6B. VEH = vehicle control; TNF = TNF-alpha treatment (30 ng/ml).

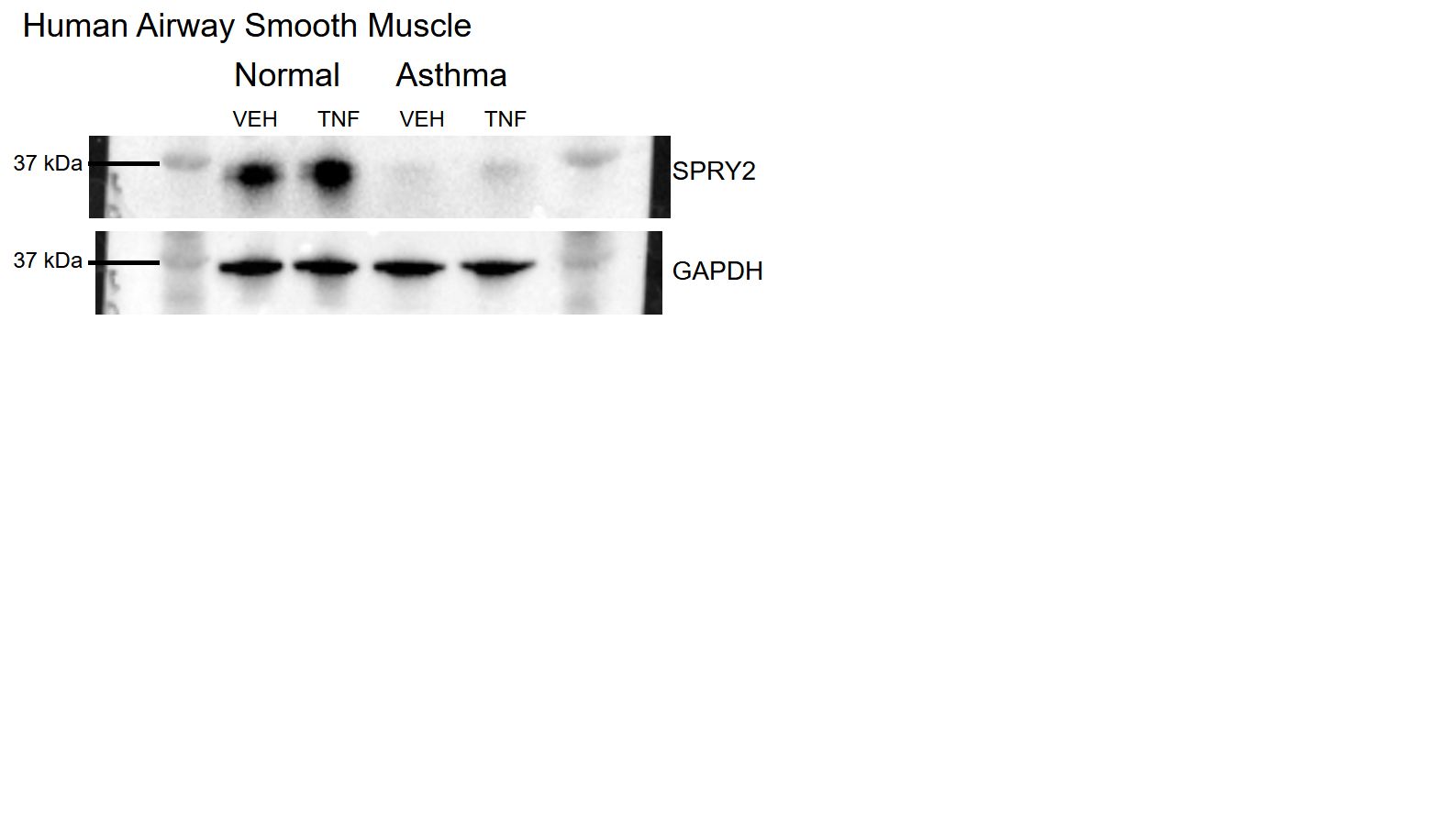

Figure E9 – *SPRY2* knockdown in primary SAECs from one donor using CRISPR-Cas9 demonstrated reduced *SPRY2* in cells nucleofected with targeting guide RNA (*SPRY2*ko) compared with non-targeting control guide RNA (NC). *SPRY2*ko VEH (vehicle/control-treated) cells exhibited reduced phosphorylated ERK1/2 and increased ERK1/2 expression. Lamin B1 served as the loading control for vehicle treated cells. Lamin B1 is cleaved by acute WSP exposure, such as in the case of the 3-hour WSP exposure results depicted below, but WSP exposure did not significantly affect ERK1/2 phosphorylation in this model.

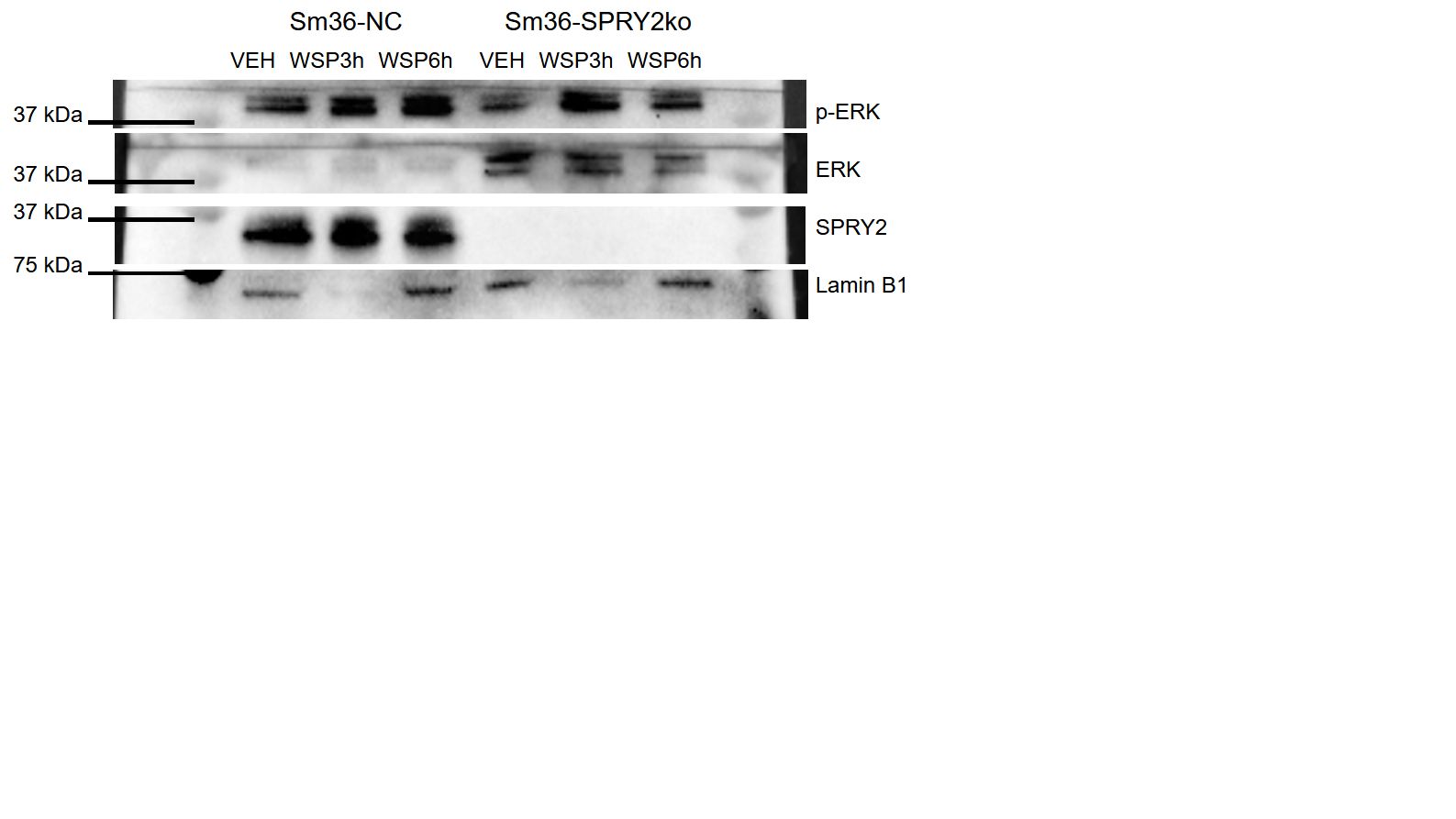
